# Connectome-wide mega-analysis reveals robust patterns of atypical functional connectivity in autism

**DOI:** 10.1101/2022.01.09.22268936

**Authors:** Iva Ilioska, Marianne Oldehinkel, Alberto Llera, Sidhant Chopra, Tristan Looden, Roselyne Chauvin, Daan Van Rooij, Dorothea L. Floris, Julian Tillmann, Carolin Moessnang, Tobias Banaschewski, Rosemary J. Holt, Eva Loth, Tony Charman, Declan G. M. Murphy, Christine Ecker, Maarten Mennes, Christian F. Beckmann, Alex Fornito, Jan K. Buitelaar, the EU-AIMS LEAP group

## Abstract

Neuroimaging studies on functional connectivity (FC) in autism have been hampered by small sample sizes and inconsistent findings with regard to whether connectivity is increased or decreased in individuals with autism, whether these alterations affect focal systems or reflect a brain-wide pattern, and whether these are age- and/or sex-dependent. The study included resting-state fMRI and clinical data from the LEAP and the ABIDE I and II initiatives, of 1824 (796 with autism) participants with age range 5-58 years. Between-group differences in FC were assessed, and associations between FC and clinical symptom ratings were investigated through canonical correlation analysis.

Autism was associated with a brain-wide pattern of hypo- and hyperconnectivity. Hypoconnectivity predominantly affected sensory and higher-order attentional networks and correlated with social impairments, restrictive and repetitive behavior (RRB), and sensory processing. Hyperconnectivity was observed primarily between the default mode network (DMN) and the rest of the brain, and between cortical and subcortical systems. This pattern was strongly associated with social impairments and sensory processing. Interactions between diagnosis and age or sex were not statistically significant.

The FC alterations observed in this study, which primarily involve hypoconnectivity of primary sensory and attention networks and hyperconnectivity of the DMN and subcortex with the rest of the brain, do not appear to be age or sex-dependent and correlate with clinical dimensions of social difficulties, RRBs, and alterations in sensory processing. These findings suggest that the observed connectivity alterations are stable, trait-like features of autism that are related to the three main symptom domains.

## INTRODUCTION

Autism spectrum disorder (henceforth autism or ASD) is a lifelong early-onset neurodevelopmental condition characterized by social-communication difficulties, restricted and repetitive behaviors, and atypical sensory processing (1). The prevalence of autism is high, with 1-2% of all people worldwide affected (2). The absence of effective treatments underscores the need to better understand the neural underpinnings of the condition. Over the last two decades, the research community has shifted its view from considering autism as a disorder of isolated brain regions to investigating the hypothesis that autism is associated with atypical interactions across distributed brain networks (3, 4).

Through advances in different network mapping techniques (5-8), recent years have seen an increasing focus on trying to understand how distinct aspects of brain connectivity are altered in individuals with autism. Early work in 2004 examining functional connectivity (FC) in autism reported decreased FC (hypoconnectivity) relative to neurotypical (NT) controls, in individuals with autism during the performance of a sentence comprehension task (9).

Several subsequent rs-fMRI studies also found decreased FC in autism (10-12). In contrast, other rs-fMRI studies have reported increased FC (hyperconnectivity) in autism, including FC between primary sensory and subcortical networks (13), and within the primary motor cortex (14), ventral attention, default mode network (DMN), and visual networks (15). Later studies, published from 2013 to 2019, put forward an even more complex picture, reporting both hyperconnectivity and hypoconnectivity, within and between various brain networks (15-18). Accordingly, while many rs-fMRI studies have reported altered FC in autism, these findings are inconsistent with respect to the specific networks implicated and whether these networks show, relative to neurotypicals, increased FC, decreased FC, or a combination of both (19).

Several factors may have contributed to these inconsistencies.

First, most studies have relied on small sample sizes, typically <70 per group (19), which can inflate the effect sizes and decrease the reproducibility of results (20). Multi-site consortia such as the Autism Brain Imaging Data Exchange (ABIDE) (21), EU-AIMS Longitudinal European Autism Project (LEAP) (22), and ENIGMA (23) have been established to overcome such sample size limitations. Second, many studies differ in terms of whether and how they account for head motion (24-26). Increased head motion can lead to artefactual findings of decreased long-distance FC and increased short-distance FC (27, 28), a pattern that has also been hypothesized in autism (29). Third, inconsistent results may also be driven by the different network mapping techniques used. Many studies either use seed-based connectivity, which only allows for investigating a limited number of brain regions (19), or spatial independent component analysis (ICA), which has a limited capacity to identify the specific connections between brain regions that may be altered. Finally, the age range of studied samples shows considerable variation, which may affect the results. For example, Uddin and colleagues identified a pattern of hyperconnectivity in children and hypoconnectivity in adolescents and adults across several studies (30).

In this study, we aim to overcome these limitations by combining data from three of the largest initiatives to date: EU-AIMS (22), ABIDE I, and ABIDE II (21). Following rigorous quality control and consideration of motion-related contamination, we combine mega-analytic (i.e., data pooling) methods with comprehensive whole-brain network mapping and analysis techniques analogous to cluster-based inference in fMRI (31) to generate connectome-wide maps of altered FC across 75,855 different connections linking 390 brain regions. Importantly, our total sample spans ages 5 to 58 years, allowing us to investigate the age dependence of any autism-related changes in FC. We use canonical correlation analysis (CCA) to relate FC alterations within autism to distinct dimensions of clinical symptomatology. Critically, we replicate the results obtained with the whole data in the EU-AIMS LEAP and ABIDE1&2 datasets independently.

Our primary goal is to test the hypothesis that autism is associated with both hypo- and hyperconnectivity, and to characterize relationships between functional connectivity alterations and clinical scores covering the three main symptom domains of autism; namely, social difficulties, restricted and repetitive behavior, and sensory processing. Our unique approach thus allows us to comprehensively evaluate the network-specificity and polarity of FC alterations in autism, their age-dependence, and their clinical correlates.

## METHODS AND MATERIALS

### Participants

We combined data from three large datasets: EU-AIMS Longitudinal European Autism Project (LEAP) (22) https://www.eu-aims.eu/ and https://www.aims-2-trials.eu/, and Autism Brain Imaging Data Exchange, ABIDE-I and ABIDE-II (21) http://fcon_1000.projects.nitrc.org/indi/abide/, see Supplement Section 1.

Our final sample for analysis included 796 individuals with autism (141 females; age-range: 5-58) and 1028 NT individuals (256 females; age-range: 5-56). Details about our exclusion criteria are provided in Supplement Section 2. The clinical and demographic characteristics of included participants are listed in Table 1. For age distributions histogram see Figure S15 in the supplement.

**Table 1.**
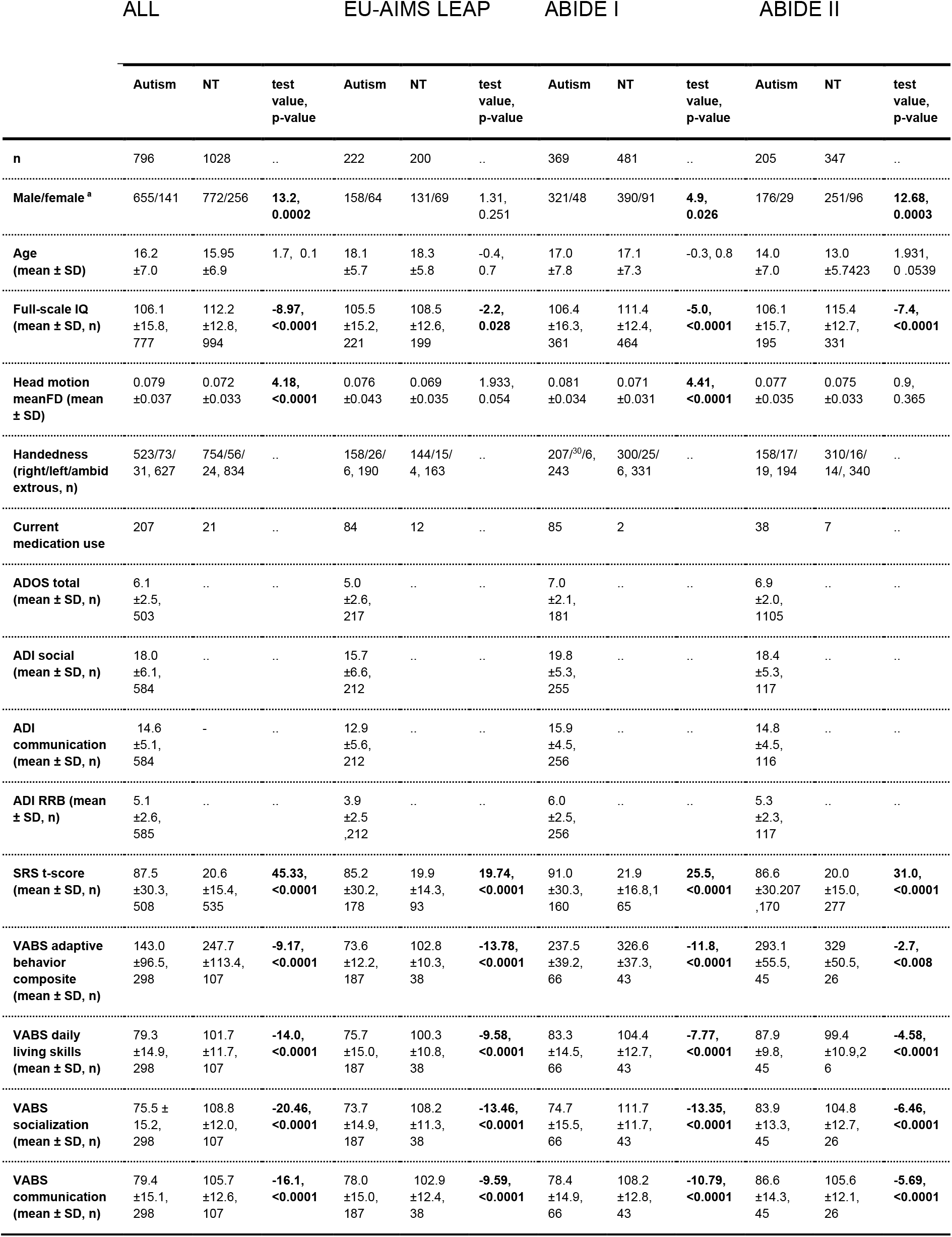
Demographic and clinical information of included participants. ADOS, Autism Diagnostic Observation Schedule; ADI-R, Autism Diagnostic Interview-Revised; autism, autism spectrum disorder; NT, neurotypicals; FD, framewise displacement; RRB, Restrictive Interests and Repetitive Behavior; SRS-2, Social Responsiveness Scale Second Edition; VABS, Vineland Adaptive Behavior Scales; “-”, not applicable; FD, framewise displacement; SD, standard

|  | ALL |  |  | EU-AIMS LEAP |  |  | ABIDE I |  |  | ABIDE II |  |  |
| --- | --- | --- | --- | --- | --- | --- | --- | --- | --- | --- | --- | --- |
|  | Autism | NT | test value, p-value | Autism | NT | test value, p-value | Autism | NT | test value, p-value | Autism | NT | test value, p-value |
| n | 796 | 1028 | .. | 222 | 200 | .. | 369 | 481 | .. | 205 | 347 | .. |
| Male/female * | 655/141 | 772/256 | **13.2, 0.0002** | 158/64 | 131/69 | 1.31, 0.251 | 321/48 | 390/91 | **4.9, 0.026** | 176/29 | 251/96 | **12.68, 0.0003** |
| Age (mean ± SD) | 16.2 ±7.0 | 15.95 ±6.9 | 1.7, 0.1 | 18.1 ±5.7 | 18.3 ±5.8 | -0.4, 0.7 | 17.0 ±7.8 | 17.1 ±7.3 | -0.3, 0.8 | 14.0 ±7.0 | 13.0 ±5.7423 | 1.931, 0.0539 |
| Full-scale IQ (mean ± SD, n) | 106.1 ±15.8, 777 | 112.2 ±12.8, 994 | **-8.97, <0.0001** | 105.5 ±15.2, 221 | 108.5 ±12.6, 199 | **-2.2, 0.028** | 106.4 ±16.3, 361 | 111.4 ±12.4, 464 | **-5.0, <0.0001** | 106.1 ±15.7, 195 | 115.4 ±12.7, 331 | **-7.4, <0.0001** |
| Head motion meanFD (mean ± SD) | 0.079 ±0.037 | 0.072 ±0.033 | **4.18, <0.0001** | 0.076 ±0.043 | 0.069 ±0.035 | 1.933, 0.054 | 0.081 ±0.034 | 0.071 ±0.031 | **4.41, <0.0001** | 0.077 ±0.035 | 0.075 ±0.033 | 0.9, 0.365 |
| Handedness (right/left/ambidextrous, n) | 523/73/31, 627 | 754/56/24, 834 | .. | 158/26/6, 190 | 144/15/4, 163 | .. | 207/<sup>30</sup>6, 243 | 300/25/6, 331 | .. | 158/17/19, 194 | 310/16/14/, 340 | .. |
| Current medication use | 207 | 21 | .. | 84 | 12 | .. | 85 | 2 | .. | 38 | 7 | .. |
| ADOS total (mean ± SD, n) | 6.1 ±2.5, 503 | .. | .. | 5.0 ±2.6, 217 | .. | .. | 7.0 ±2.1, 181 | .. | .. | 6.9 ±2.0, 1105 | .. | .. |
| ADI social (mean ± SD, n) | 18.0 ±6.1, 584 | .. | .. | 15.7 ±6.6, 212 | .. | .. | 19.8 ±5.3, 255 | .. | .. | 18.4 ±5.3, 117 | .. | .. |
| ADI communication (mean ± SD, n) | 14.6 ±5.1, 584 | - | .. | 12.9 ±5.6, 212 | .. | .. | 15.9 ±4.5, 256 | .. | .. | 14.8 ±4.5, 116 | .. | .. |
| ADI RRB (mean ± SD, n) | 5.1 ±2.6, 585 | .. | .. | 3.9 ±2.5, 212 | .. | .. | 6.0 ±2.5, 256 | .. | .. | 5.3 ±2.3, 117 | .. | .. |
| SRS t-score (mean ± SD, n) | 87.5 ±30.3, 508 | 20.6 ±15.4, 535 | **45.33, <0.0001** | 85.2 ±30.2, 178 | 19.9 ±14.3, 93 | **19.74, <0.0001** | 91.0 ±30.3, 160 | 21.9 ±16.8, 165 | **25.5, <0.0001** | 86.6 ±30.207, 170 | 20.0 ±15.0, 277 | **31.0, <0.0001** |
| VABS adaptive behavior composite (mean ± SD, n) | 143.0 ±96.5, 298 | 247.7 ±113.4, 107 | **-9.17, <0.0001** | 73.6 ±12.2, 187 | 102.8 ±10.3, 38 | **-13.78, <0.0001** | 237.5 ±39.2, 66 | 326.6 ±37.3, 43 | **-11.8, <0.0001** | 293.1 ±55.5, 45 | 329 ±50.5, 26 | **-2.7, <0.008** |
| VABS daily living skills (mean ± SD, n) | 79.3 ±14.9, 298 | 101.7 ±11.7, 107 | **-14.0, <0.0001** | 75.7 ±15.0, 187 | 100.3 ±10.8, 38 | **-9.58, <0.0001** | 83.3 ±14.5, 66 | 104.4 ±12.7, 43 | **-7.77, <0.0001** | 87.9 ±9.8, 45 | 99.4 ±10.9, 26 | **-4.58, <0.0001** |
| VABS socialization (mean ± SD, n) | 75.5 ±15.2, 298 | 108.8 ±12.0, 107 | **-20.46, <0.0001** | 73.7 ±14.9, 187 | 108.2 ±11.3, 38 | **-13.46, <0.0001** | 74.7 ±15.5, 66 | 111.7 ±11.7, 43 | **-13.35, <0.0001** | 83.9 ±13.3, 45 | 104.8 ±12.7, 26 | **-6.46, <0.0001** |
| VABS communication (mean ± SD, n) | 79.4 ±15.1, 298 | 105.7 ±12.6, 107 | **-16.1, <0.0001** | 78.0 ±15.0, 187 | 102.9 ±12.4, 38 | **-9.59, <0.0001** | 78.4 ±14.9, 66 | 108.2 ±12.8, 43 | **-10.79, <0.0001** | 86.6 ±14.3, 45 | 105.6 ±12.1, 26 | **-5.69, <0.0001** |

### Clinical diagnosis

Individuals with autism in the EU-AIMS LEAP dataset had an existing clinical diagnosis of autism according to the criteria of DSM-IV, DSM-5, or ICD-10 with the majority also meeting the autism criteria on Autism Diagnostic Interview-Revised (ADI-R) (32) and/or Autism Diagnostic Observation Schedule 2 (ADOS 2) (33); see Charman et al. 2017 (34). Each site within ABIDE followed their individual diagnostic procedures for establishing an autism diagnosis, although the majority of the sites used the Autism Diagnostic Observation Schedule (35) and/or Autism Diagnostic Interview-Revised (32). Neurotypical controls are individuals that reported no psychiatric diagnosis.

### MRI acquisition and Pre-processing

Resting-state fMRI and structural scans were obtained using 3T MRI scanners at 32 scanning sites (Supplement Section 3). The Supplement also contains details on denoising efficacy (Supplement Section 6) and the sensitivity of our findings to different preprocessing procedures (Supplement Section 9). ComBat (36) was used to remove the confounding effects of scanning site from the fMRI data (Supplement Section 4).

### Mapping functional connectivity

To investigate autism-related FC alterations across the brain, we applied the well-validated Schaefer parcellation to divide the cortex of each participant into 400 functional regions of interest (ROIs) (37), combined with 15 subcortical ROIs from the Harvard-Oxford subcortical atlas (38) We extracted the mean time series from each ROI and computed the Pearson correlation between every pair of time series. The values within each participants’ FC matrix were normalized using Gaussian-gamma mixture modeling (39) (Supplement Section 5).

### Analysis of group differences

Through a mega-analytic approach (the pooling of raw data acquired at multiple sites), we mapped connectome-wide differences between autistic and neurotypical individuals using the Network Based Statistic (NBS) (31) (software available at www.nitrc.org/projects/nbs/). To assess FC group differences, a general linear model was fitted to each edge, including diagnosis, age, sex, and mean framewise displacement (FD) as covariates. We additionally examined interactions between diagnosis and age and diagnosis and sex. For details about the NBS procedure, see Supplement Section 7. We report results at a component-wide, familywise error-corrected threshold of p_fwe_<.001, following the application of primary component thresholds corresponding to *p* < {.05, .01, and .001} (40).

We used the Yeo 7-network parcellation to assign cortical ROIs to well-known resting-state networks (41). These assignments allowed us to quantify the number of edges identified by the NBS as showing an effect of interest within and between each of the networks (N_raw_). As the number of ROIs and potential connections within and between networks differs, we calculated the proportion of altered edges corrected by the total amount of possible edges within or between the particular networks (N_norm_). Thus, raw counts allow us to identify networks that contribute strongly to group differences in absolute terms; normalized counts allow us to identify strong contributions relative to network size. We also calculated the degree centrality of each region, defined as the number of connections attached to each node in the NBS subnetwork, thus identifying specific regions heavily involved in connectivity differences. To further demonstrate the robustness and replicability of our findings we conducted the full FC mapping and analysis of group differences separately in the LEAP and ABIDE 1&2 data.

### Brain-behavior correlations

We used CCA to relate autism-related FC connectivity to symptom severity. In our analyses, one set of variables included measures of behavior and symptom severity, and the other set of measures included principal components of the FC estimates for edges identified as showing atypical connectivity in our NBS analysis. We interpreted the results by directly correlating the edge-level measures with the canonical variates identified by the CCA, allowing us to determine the contribution of each edge to the canonical variate, providing a clearly interpretable brain-behavior relationship.

We conducted two CCA analyses. In the first CCA, we investigated associations with a diverse set of behavioral variables, including the ADOS social affect, ADOS restricted and repetitive behaviors (RRB), ADI RRB, ADI communication, ADI Social, SRS-2, and VABS daily living skills, VABS communication, and VABS social relationships. These measures were collected across the three datasets for a subsample of 232 individuals with autism. Given that the SSP scores were only available for a smaller sample of 125 individuals with autism and 78 controls from the LEAP cohort, we conducted a separate, CCA examining FC relationships with the subscales of the SSP (42). The behavioral measures are detailed in Supplement Section 8. We further validated our CCA findings using leave-one-site-out cross-validation as explained in Supplement Section 13.

## RESULTS

### Atypical FC in autism

At the primary component-forming threshold of *p*<0.05, the NT>ASD contrast revealed hypoconnectivity in autism that encompassed 13,001 edges (17.1% of the total number of edges) linking 376 brain regions (*p*_*fwe*_<0.001; Figure 1A-C). The ASD>NT contrast revealed hyperconnectivity in autism, including 16,767 edges (22% of the total number of edges; *p*_*fwe*_<0.001) and linking all 390 brain regions (Figure 1D-F). Thus, nearly all brain regions were implicated in both the hyper- and hypoconnectivity findings in ASD.

**1.**
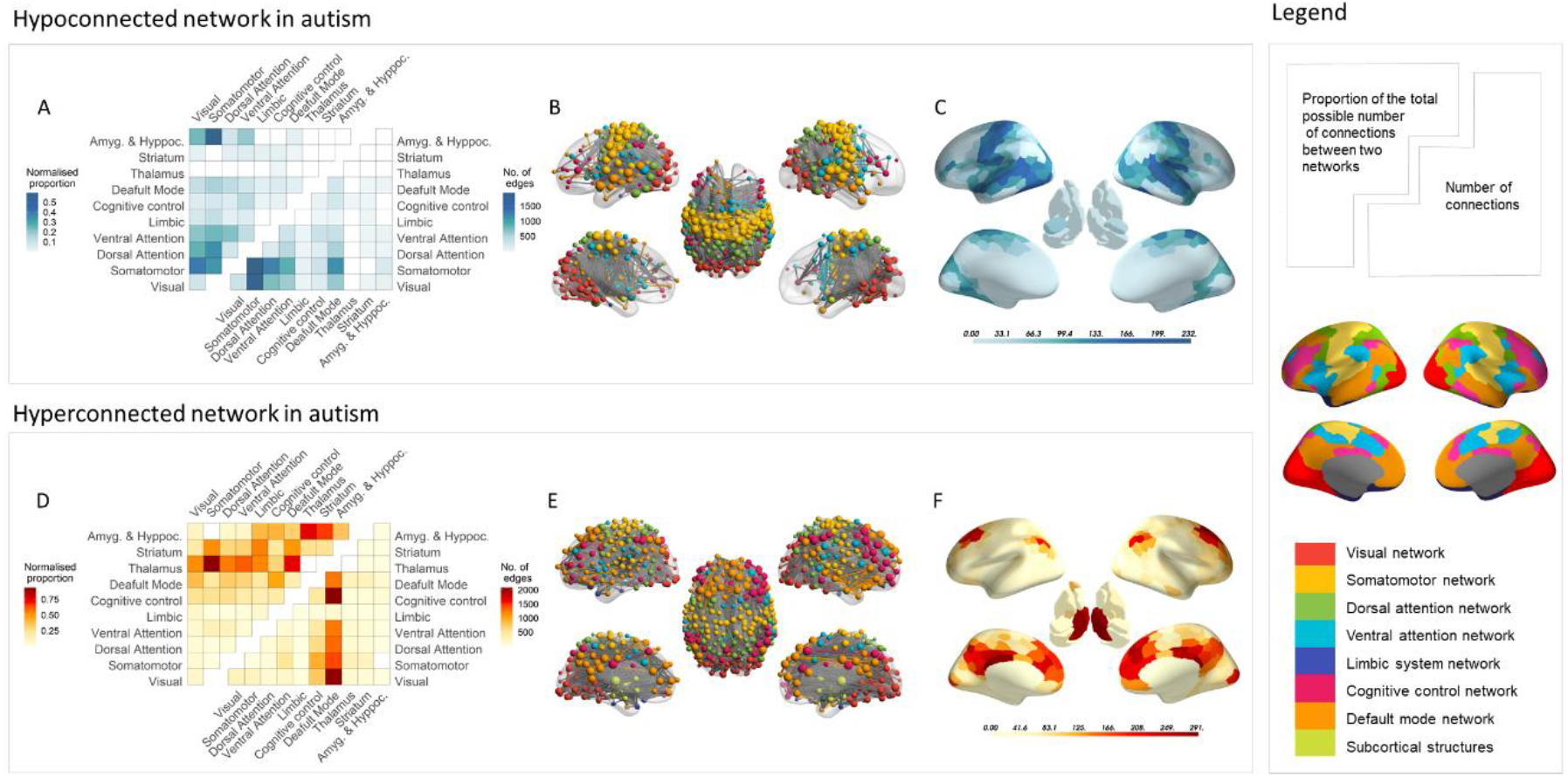
Networks of altered functional connectivity in ASD. A) Matrix indicating the number of decreased connections (TD>ASD contrast) between and within networks at the threshold corresponding to *p*<0.05; As indicated in the legend, the upper triangle of the matrix shows the proportion of decreased connections falling within and between each of the networks, normalized by the total number of possible connections within or between the corresponding networks; the lower triangle shows the raw count of edges. (B) Anatomical projection of the edges showing significant hypoconnectivity in ASD. C) Cortical and subcortical maps showing the degree centrality of each region within the NBS-identified hypoconnectivity network in ASD; D) Matrix indicating the number of increased connections (ASD>TD contrast) between and within networks at the threshold corresponding to *p*<0.05; As indicated in the legend, the upper triangle of the matrix shows the proportion of increased connections falling within and between each of the networks, normalized by the total number of possible connections within or between the corresponding networks; the lower triangle shows the raw count of edges; E) Anatomical projection of the edges showing significant hyperconnectivity in ASD; F) Cortical and subcortical maps showing the degree centrality of each region within the NBS-identified hyperconnectivity network in ASD

The highest proportion of hypoconnected edges was present between the visual and somatomotor networks (N_norm_=0.4; N_raw_=1862), followed by within-network connectivity of the somatomotor network (N_norm_=0.33; N_raw_=1924), and between-network connectivity of the somatomotor network and the dorsal attention (N_norm_=0.35; N_raw_=1225) and salience systems (N_norm_=0.24; N_raw_=875; 1A;1B). FC between the lateral temporal regions of the DMN and sensory and attention networks also featured prominently (Figure 1B). Accounting for the differences in network size, further implicated connectivity between the amygdala and the somatomotor (N_norm_=0.59, N_raw_=180) and visual networks (N_norm_=.24; N_raw_=59; Figure 1A and 1C).

Atypical hyperconnectivity was predominantly found between the DMN and the rest of the brain, particularly the cognitive control network (N_norm_=0.46; N_raw_=2069; Figures 1D; 1E). When accounting for differences in network size, atypical FC between the thalamus and the rest of the brain, and between medial temporal areas and thalamus and striatum, also featured prominently (Figure 1D; 1E).

At the primary component-forming thresholds of *p*<0.01 and *p*<0.001, the affected networks were sparser, but a similar pattern of both hypoconnectivity and hyperconnectivity was evident (Supplement Figure S5). Results were similar when using global signal regression, and when including IQ as a covariate (Supplement Sections 9 and 10), supporting the robustness of our findings. Our conclusions remain unchanged in light of the sensitivity analyses for sex, medication, and ADHD comorbidity and analysis including subjects with IQ<70, which can be found in Supplement Section 10.

### Age- and sex-dependent FC changes in autism

We identified marginal evidence for an age-by-diagnosis interaction for the ASD>NT contrast (*p*_*fwe*_=0.08) at the primary threshold of *p*<0.05 (Supplement Section 12). No significant sex-by-diagnosis interactions were identified.

### Split-sample analysis

To further demonstrate the robustness and replicability of our findings we conducted the full FC mapping and analysis of group differences separately in the LEAP and ABIDE 1&2 data. Our results of an atypical pattern of hyper- and hypoconnectivity were largely consistent when analyzing the LEAP and ABIDE 1&2 datasets separately (Figure S11). The network counts of the two results were highly correlated (hypoconnectivity: r=0.93, p=0; hyperconnectivity: r=0.82, p=0). Specifically, the separate analyses of both cohorts revealed consistent evidence for hyperconnectivity between the DMN with the rest of the brain and hyperconnectivity between the thalamus and the rest of the brain. The observed hypoconnectivity within the somatomotor network and between the primary somatosensory, visual, and attentional networks was also consistent across the two datasets (Supplement Section 11).

### Brain-behavior correlations

We next used CCA to evaluate brain-behavior correlations between ten behavioral measures and 48 PCs explaining 50% of the FC variance. This analysis revealed two significant CVs (CV1: r=0.72, *p*<0.0001; CV2: r=0.65, *p*<0.0001; see Figures 2A and 2E).

**2.**
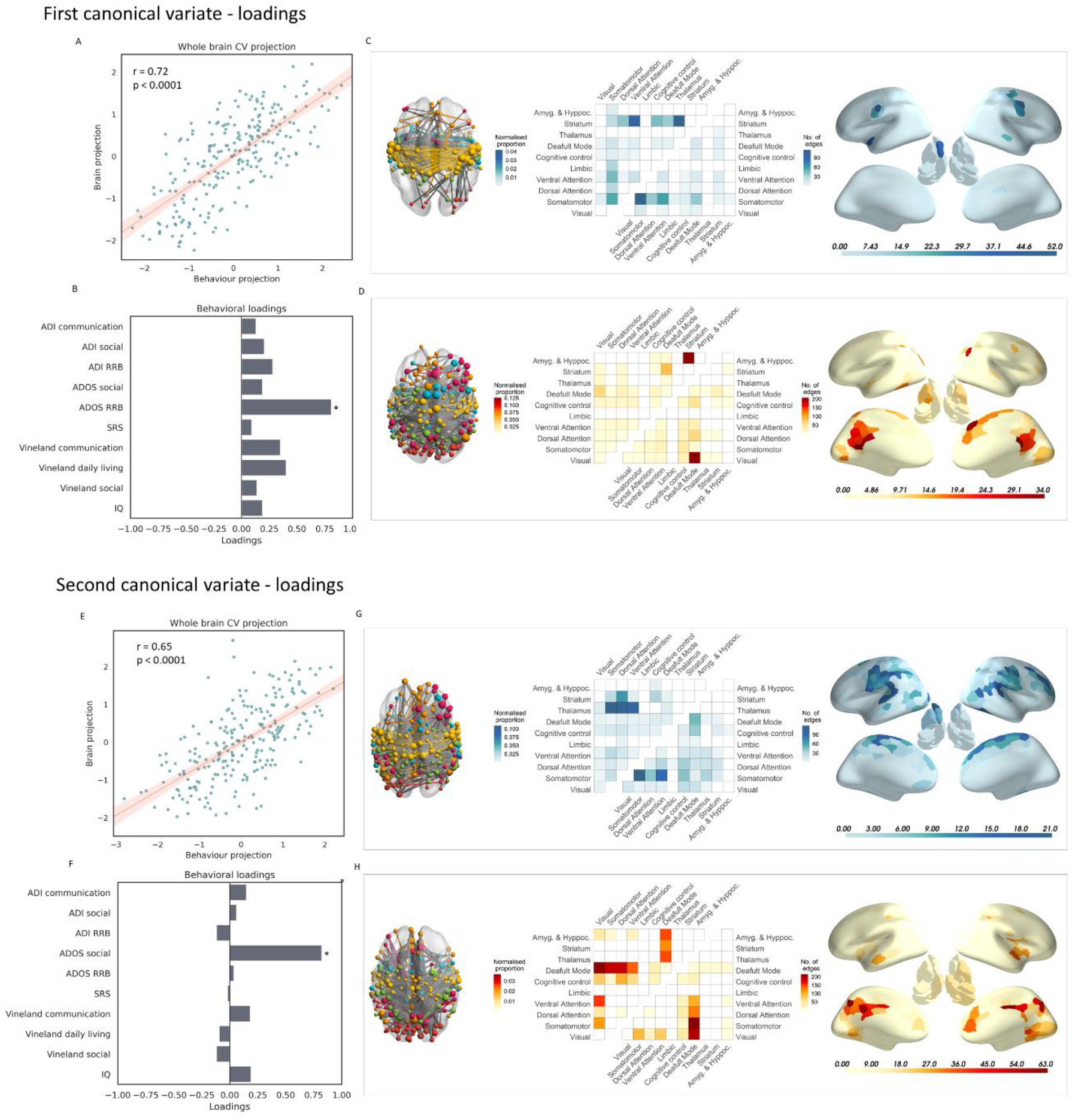
Brain-behavior correlations (Social, RRB, adaptive behavior, and IQ) A) Correlation of the first pair of canonical variates (CV1: r=0.72, p<0.0001) resulting from the canonical correlation analysis linking the behavioral scores to the principal components accounting for 50% of the FC-hypoconnectivity combined with FC-hyperconnectivity variance; B) Structural coefficients, also known as loadings of the behavioral scores with the first CV; C) Significant negative correlations of FC with CV1, :p<0.05, characterized at the edge, network and region level; D) Significant positive correlations of FC with CV1: p<0.05, characterized at the edge, network and region level; E). Correlation of the second pair of canonical variates (CV2: r=0.65, p<0.0001); F) Structural coefficients, also known as loadings of the behavioral scores with the second CV; G) Significant negative correlations of FC with CV2:p<0.05, characterized at the edge, network and region level; H) Significant positive correlations of FC with CV2:p <0.05, characterized at the edge, network and region level.

The first CV exhibited a very strong association with the restricted interests and repetitive behavior scale of the ADOS (ADOS RRB; loading=0.82; Figure 2B). Analysis of the FC loadings revealed that higher RRB scores were associated with lower FC within the somatomotor network and between this network and the dorsal and ventral attention systems (Figure 2C); higher FC between the DMN and visual network; and higher FC between the dorsal attention and cognitive control networks (Figure 2D). Normalized proportions additionally revealed a contribution of lower striatal connectivity with nearly all other systems, when accounting for network size. Degree centrality analysis identified the left nucleus accumbens and the somatomotor cortex as regions with many connections where lower FC was associated with RRBs, and posterior cingulate and medial frontal cortex, as regions where higher FC was associated with RRBs in ASD (Figure 2D).

The second CV exhibited a strong association with the ADOS Social Affect score (loading=0.82; see Figure 2F). Higher ADOS Social Affect scores were associated with higher FC between the DMN and the rest of the brain, particularly the visual network (Figure 2H). Higher FC of the cuneus, lingual gyrus, and posterior cingulate cortex was implicated in this relationship (Figure 2H). Greater social impairment was also associated with lower connectivity mainly within the somatomotor network and between the somatomotor network and higher-order attentional networks; and between the thalamus and somatomotor and attentional networks (Figure 2G).

In our second CCA analysis, we evaluated brain-behavior correlations between the six SSP subscale scores and 44 PCs explaining 50% of the variance across both controls and individuals with autism. This analysis revealed one significant CV (CV1: r=0.63, p<0.0001; see Figure 3A). The loadings of CV1 revealed a significant association with sensation seeking (loading = 0.85), visual/auditory sensitivity (loading=0.72) and movement sensitivity (loading=0.74; Figure 3B). Greater sensory difficulties were associated with lower FC between the visual and somatomotor network and between the amygdala and somatomotor network (Figure 3C); and higher FC between somatomotor and cognitive control network and the DMN and nearly all other networks, especially the ventral attention, and cognitive control network. Higher within network connectivity of the DMN was also associated with atypical sensory processing. Normalized proportions revealed that sensory difficulties were related to higher FC between the thalamus and the somatomotor network (Figure 3D).

**3.**
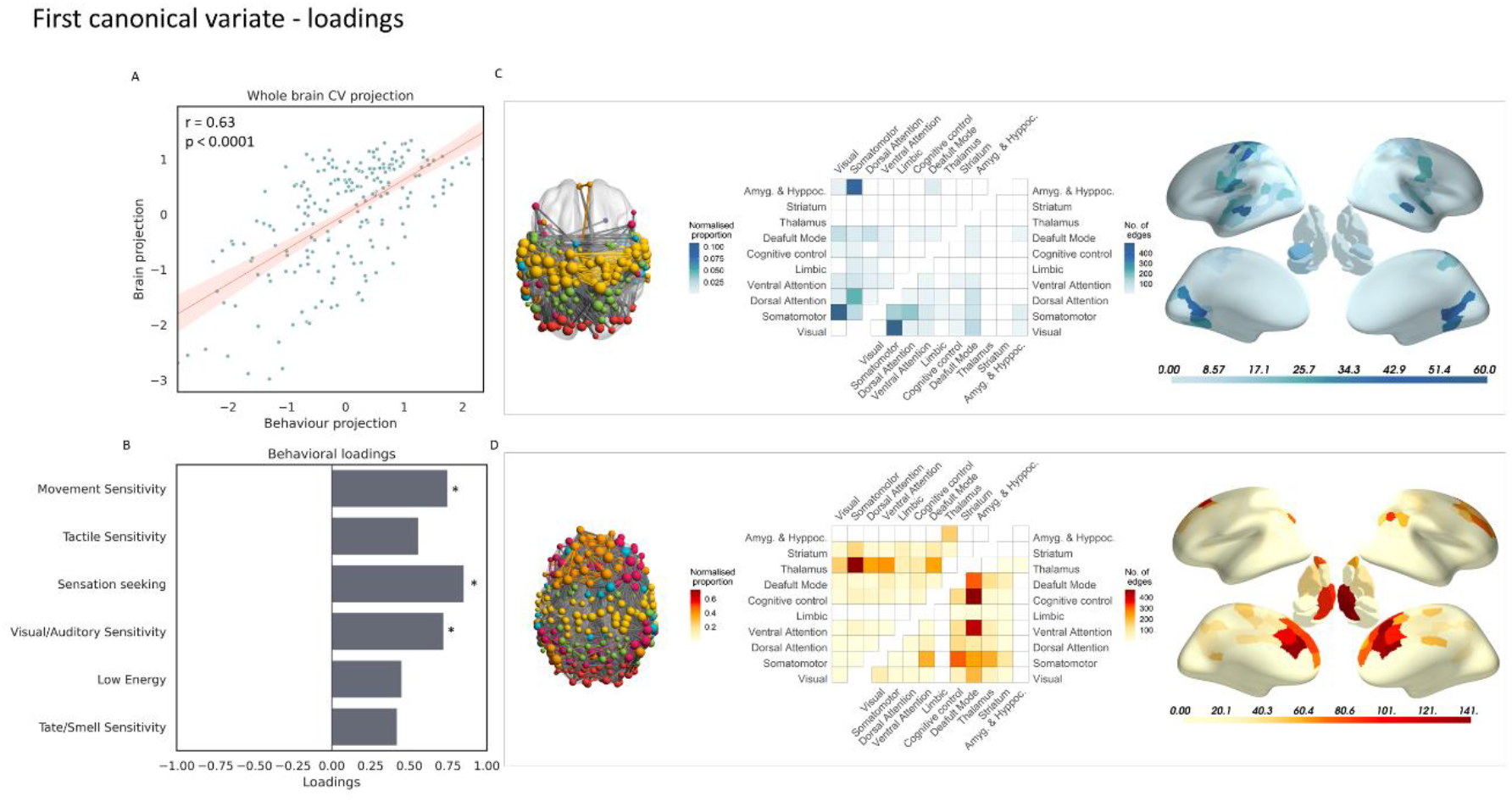
Brain-behavior correlations (Short Sensory Profile subscales) A) Correlation of the first pair of canonical variates (CV1: r=0.63, p<0.0001) resulting from the canonical correlation analysis linking the behavioral SSP scores to the principal components accounting for 50% of the FC-hypoconnectivity combined with FC-hyperconnectivity variance; B) Structural coefficients, also known as loadings of the behavioral scores with the first CV; C) Significant negative correlations of FC with CV1:p<0.05, characterized at the edge, network and region level; D) Significant positive correlations of FC with CV1: p<0.05, characterized at the edge, network, and region level

We additionally conducted leave-one-site-out validation analyses, which yielded consistent findings (Supplement Figures S13 and S14; Tables S11 and S12).

## DISCUSSION

Using a comprehensive connectome-wide mega-analysis, we report that autism is associated with complex, widely distributed differences in functional connectivity that encompass the entire brain. We demonstrate the robustness of these findings by successful replication in separate analyses of the LEAP and ABIDE1&2 datasets. Reduced FC is predominantly found within the somatomotor network and between the sensory – motor processing networks and higher-order attentional systems. Increased FC is predominantly found between the DMN, subcortex, and the rest of the brain. These patterns show strong associations with sensory processing difficulties and distinct associations with clinical variables, such that lower connectivity of sensorimotor and attentional networks is associated with social difficulties and restricted interests and repetitive behaviors, whereas higher FC between the DMN and other systems is mainly associated with social impairments. We found marginal evidence for age-dependent FC changes and no interaction between diagnosis and sex. Our findings thus indicate that autism is characterized by a relatively stable pattern of widely distributed increases and decreases in functional connectivity that are related to clinical symptoms of the disorder.

### Hypoconnectivity in autism

Our results indicate that hypoconnectivity in autism is most prevalent for functional connections within the somatomotor network and connections linking the visual and somatomotor network to each other and to attentional networks. Several studies have previously reported atypical sensory processing and visual-motor integration in autism (14, 43-47). Our finding of hypoconnectivity between the visual and somatomotor network is furthermore consistent with recent work (16) showing lower connectivity among visual association, somatosensory, and motor networks in autism.

The CCA revealed that while lower connectivity within the somatomotor network and between the somatomotor network and the attentional networks is related to RRBs and social impairments, lower connectivity between the somatomotor network and the visual network is related to three subscales of sensory processing (SSP). The link between altered sensory processing and social impairments and RRBs in autism is supported by the previous literature (48-50). Lower connectivity of the somatomotor cortex has been associated with atypical primary sensory processing, which might potentially hamper the ability to filter relevant information and recruit a suitable motor response, leading to reduced integration of the somatomotor networks with the salience and dorsal attention networks (48). RRBs have been suggested to provide a way to manage atypically regulated sensory stimulation by means of reducing overstimulation or creating stimulation in cases of stimulation seeking (48).

### Hyperconnectivity in autism

Our results further revealed prominent hyperconnectivity within the DMN and between the DMN and the rest of the brain, particularly with the cognitive control and visual networks in autistic individuals. Thalamocortical connectivity and connectivity between medial temporal and striato-thalamic systems also showed prominent connectivity increases relative to network size. The CCA indicated that the observed pattern of hyperconnectivity, particularly the between-network connectivity of the DMN, was linked to sensory processing difficulties and social impairments (ADOS-SA). FC between the DMN and the visual network was also strongly associated with restrictive and repetitive behaviors and social impairments, whereas FC within the DMN, between the DMN and the ventral attention and frontoparietal networks, was strongly linked to atypical sensory processing.

Our findings accord with many prior rs-fMRI studies implicating disrupted connectivity of the DMN (15, 51, 52), and disrupted connectivity between the DMN, cognitive control, and other networks in autism (53, 54). Our findings are also consistent with previous work showing increased subcortical-cortical connectivity in autism (13, 21, 55). The observed associations with social impairments suggest that the general pattern of hyperconnectivity in autism may limit dynamic interactions between networks, potentially leading to decreased behavioral flexibility and thus difficulties in navigating dynamic real-world social situations (15).

### The effects of age and sex

We found marginal evidence of an interaction between age and diagnosis, but this result did not survive our significance criteria. We, therefore, did not replicate previously reported findings suggesting developmental effects in autism (30). This may be the result of possible heterogeneous developmental trajectories in the group of individuals with autism and demonstrates an absence of a robust general developmental trajectory distinguishing the autistic from neurotypical development after the age of 6. We did not observe a significant sex-by-diagnosis interaction, suggesting limited evidence of sexually distinct patterns of altered functional connectivity in autism (56).

### Limitations

Our analysis did not include the cerebellum due to poor scan coverage in many participants. Although our mega-analysis allowed us to examine participants across a relatively broad age range, most participants were under 25 years of age. This is consistent with a general trend in the literature to focus on younger samples. As such, it is still unclear how ASD affects the brain beyond the third and fourth decades of life. It should also be noted that our sample sex ratio was imbalanced between the autism and control groups, reflecting the higher proportion of males compared to females diagnosed with autism in the general population. An important way forward for the field will be to target individuals with autism who are female and particularly those over 30 years of age. Finally, even though our study shows many significant case-control differences in functional connectivity at the group level, these results should be viewed in light of the considerable heterogeneity present across individuals with autism. As such, future work will focus on characterizing heterogeneity in autism and defining autism connectivity subtypes.

### Conclusion

Our connectome-wide mega-analysis identified a widespread and robust pattern of altered functional connectivity in autism, characterized by prominent hypoconnectivity within sensory processing networks, and between these sensory networks and attentional systems, in addition to hyperconnectivity of the DMN and thalamus, with the rest of the brain. These connectivity differences were associated with distinct clinical dimensions and show limited evidence of age or sex-dependence, suggesting that they are a stable, trait-like feature of autism. Additionally, our findings replicate in two independent sub-samples. These findings do not only reveal a robust pattern of connectivity alterations, but also confirm the importance of both the DMN and sensorimotor systems in the pathophysiology of autism, and highlight their clinical relevance for understanding the symptoms of autism. This work paves the way for further work on developing connectivity biomarkers of autism and subtyping of autism that will inform clinical practice.

## Data Availability

Data from EU-AIMS LEAP is available upon application and approval to the EU-AIMS LEAP committee (https://www.eu-aims.eu/). 
Data from the ABIDE initiative is publicly available (https://fcon_1000.projects.nitrc.org/indi/abide/)  

## Funding

This work is primarily supported by the EU-AIMS consortium (European Autism Interventions), which receives support from Innovative Medicines Initiative Joint Undertaking Grant No.115300, the resources of which are composed of financial contributions from the European Union’s Seventh Framework Programme (Grant No. FP7/2007-2013), from the European Federation of Pharmaceutical Industries and Associations companies’ in-kind contributions; and by the AIMS-2-TRIALS consortium (Autism Innovative Medicine Studies-2-Trials), which has received funding from the Innovative Medicines Initiative 2 Joint Undertaking under grant agreement No. 777394, and this Joint Undertaking receives support from the European Union’s Horizon 2020 research and innovation programme and EFPIA and AUTISM SPEAKS, Autistica, SFARI. The views expressed are those of the author(s) and not necessarily those of the IMI 2JU. II is supported by an internal grant by Radboudumc/DCMN (2018-2022; to JKB and AF). MO is supported by ZonMW Rubicon Grant No. 452172019. This work has been further supported by the European Community’s Horizon 2020 Programme (H2020/2014-2020) Grant Nos. 643051 (MiND; to JKB), 642996 (BRAINVIEW; to JKB) and 847818 (CANDY; to JKB and CFB). AF was supported by the Sylvia and Charles Viertel Charitable Foundation and National Health and Medical Research Council (ID: 3274306).

## Acknowledgements

The LEAP team consists of: Jumana Ahmad, Sara Ambrosino, Sarah Baumeister, Carsten Bours, Michael Brammer, Daniel Brandeis, Claudia Brogna, Yvette de Bruijn, Ineke Cornelissen, Daisy Crawley, Guillaume Dumas, Jessica Faulkner, Vincent Frouin, Pilar Garcés, David Goyard, Joerg Hipp, Rosemary Holt, Meng-Chuan Lai, Xavier Liogier D’ardhuy, Michael V. Lombardo, David J. Lythgoe, René Mandl, Andre Marquand, Maarten Mennes, Andreas Meyer-Lindenberg, Nico Mueller, Bethany Oakley, Laurence O’Dwyer, Marianne Oldehinkel, Gahan Pandina, Barbara Ruggeri, Amber Ruigrok, Jessica Sabet, Roberto Sacco, Antonia San José Cáceres, Emily Simonoff, Will Spooren, Roberto Toro, Heike Tost, Jack Waldman, Steve C.R. Williams, Caroline Wooldridge, and Marcel P. Zwiers

## Notes

### Competing Interest Statement

JKB has been in the past three years a consultant to / member of advisory board of / and/or speaker for Takeda/Shire, Roche, Medice, Angelini, Janssen, and Servier. He is not an employee of any of these companies, and not a stock shareholder of any of these companies. He has no other financial or material support, including expert testimony, patents, royalties. TC has served as a paid consultant to F. Hoffmann-La Roche Ltd. and Servier; and has received royalties from Sage Publications and Guilford Publications.

### Author Declarations

The study was approved by national and local ethics review boards at each of the five study sites from EU-AIMS LEAP. London-Central and Queen Square Health Research Authority Research Ethics Committee approved the studies at the Autism Research Centre, University of Cambridge, and King's College London. Radboud universitair medisch centrum Instituut Waarborging Kwaliteit en Veiligheid Commissie Mensgebonden Onderzoek Regio Arnhem-Nijmegen (Radboud University Medical Centre Institute Ensuring Quality and Safety Committee on Research Involving Human Subjects Arnhem-Nijmegen) approved the studies at the Radboud University Nijmegen Medical Centre and University Medical Centre Utrecht. UMM Universitatsmedizin Mannheim, Medizinische Ethik Kommission II (UMM University Medical Mannheim, Medical Ethics Commission II) approved the study at the Central Institute of Mental Health in Mannheim. The data from the ABIDE initiative was publicly available.

