## Supplemental material for "Connectome-wide mega-analysis reveals robust patterns of atypical functional connectivity in autism"

Supplemental content

### Section 1. Datasets

We combined data from three large datasets: EU-AIMS Longitudinal European Autism Project (LEAP): https://www.eu-aims.eu/ & https://www.aims-2-trials.eu/ and Autism Brain Imaging Data Exchange, ABIDE-I and ABIDE-II: http://fcon_1000.projects.nitrc.org/indi/abide/.

LEAP is a large multicentre European initiative aimed at the stratification and identification of biomarkers in autism (1). LEAP recruited 437 individuals with autism and 300 neurotypical (NT) individuals across five different centers. Both male and female participants, aged between 6 and 30 years, were included. All participants were assessed with the same MRI protocol and underwent a comprehensive clinical, cognitive, and MRI assessment. For further details about the LEAP study, see (1, 2).

ABIDE is a data-sharing consortium that provides access to collected rs-fMRI and structural MRI data of participants with autism and matched controls (3). As such, ABIDE consists of studies that were initially conducted at each of the individual sites and only retrospectively aggregated into one dataset. ABIDE-II is an extension of the ABIDE-I initiative. Both ABIDE efforts combined provide 2,226 unique datasets collected at 32 sites, comprising 1,060 individuals with autism and 1,166 controls aged 5-64.

### Section 2. Exclusion criteria

The EU-AIMS LEAP dataset included 617 participants for which demographic and rs-fMRI data available. We excluded participants with low-quality resting-state fMRI scans for which the preprocessing failed (N=2), and participants with an incomplete scan (N=8). To account for excessive head motion during the resting-state fMRI scan, we also excluded participants based on having at least one of the following criteria: a mean framewise-displacement (mFD)>0.25mm, more than 20% of the FDs above 0.2mm, any FDs larger than 5mm (N=123). These criteria align with the "stringent" exclusion criteria advocated in prior work (4-6).

We next excluded participants based on signal coverage across our parcellated ROIs. To this end, we first excluded brain regions with more than 30% signal dropout in more than 5% of the whole subject sample (Figure S1), after which we exclude subjects that have poor coverage (<70 %) in more than 10% of the remaining regions. No participants were excluded based on this procedure in the EU-AIMS LEAP cohort (N=0).

After excluding individuals according to the above criteria, we visually inspected carpet plots for each participant (7, 8). These plots show the full fMRI time series in matrix form, with a normalized color scale used to indicate signal fluctuations at each time point in each voxel, and are excellent for identifying gross imaging artifacts and signal contributions from brain-wide signal changes. We used these plots to further exclude an additional 17 participants with visible artifacts. All participants with IQ<70 (N=51) and participants with a structural brain abnormality (N=4) were also excluded, resulting in the inclusion of 422 subjects.

In the ABIDE I cohort, 1110 participants had available demographic and resting-state fMRI data. We excluded participants with low-quality scans for which the preprocessing failed (N=9) and participants with an incomplete scan (N=8). To account for excessive head motion during the resting-state fMRI scan, we also excluded participants based on having at least one of the following criteria: a mean framewise-displacement (mFD)>0.25mm, more than 20% of the FDs above 0.2mm, any FDs larger than 5mm (N = 173) (4-6). We excluded participants with low coverage in more than 10% of the remaining brain regions (N=20) after excluding the brain regions that had more than 30% of signal dropout. Following visual inspection of the functional carpet plots(7), we excluded participants with visible artifacts (N=8). We further excluded all participants with IQ<70 (N=22) and participants with structural brain abnormalities (N=4). This resulted in the inclusion of 852 subjects.

In the ABIDE II cohort, we included data from sites with TR>800ms, out of which 902 participants had available demographic and resting-state fMRI data. We excluded participants with an incomplete scan (N=209). To account for excessive head motion during the rs-fMRI scan, we also excluded participants based on having at least one of the following criteria: a mean framewise-displacement (mFD)>0.25mm, more than 20% of the FDs above 0.2mm, any FDs larger than 5mm (N=130) ) (4-6). No participants had low coverage in more than 10% of the remaining brain regions (N=0), after excluding the brain regions with more than 30% of signal dropout. After visual inspection of the functional data carpet plots(7), we excluded participants with visible artifacts (N=9). We further excluded all participants with IQ<70 (N=2). This resulted in the inclusion of 552 subjects.

### Section 3. MRI acquisition and Pre-processing

Within the EU-AIMS LEAP consortium, MRI data were acquired at five sites: Cambridge University, King's College London (KCL), Central Institute of Mental Health Mannheim, Radboud University Nijmegen Medical Centre, University Medical Center Utrecht, using a standardized MRI protocol. The structural scan was acquired using a magnetization prepared rapid gradient-echo (MPRAGE) sequence according to the ADNI 2 / GO protocol (http://adni.loni.usc.edu). The resting-state fMRI scan was acquired using a multi-echo planar imaging sequence (9). For details on the scanning acquisition parameters in the EU-AIMS LEAP cohort, see Table S1.

In the ABIDE I cohort, we used data from sixteen sites: California Institute of Technology (Caltech), Carnegie Melon University (CMU), Kennedy Krieger Institute (KKI), University of Leuven (Leuven), Ludwig Maximilians University Munich

(MaxMun), NYU Langone Medical Center (NYU), Olin Institute of Living at Hartford Hospital (Olin), University of Pittsburgh School of Medicine (Pitt), Social Brain Lab BCN NeuroImaging Center, University Medical Center Groningen (SBL), San Diego State University (SDSU), Stanford University (Stanford), Trinity Centre for Health Sciences (Trinity), University of California Los Angeles (UCLA), University of Michigan (UM), University of Utah School of Medicine (USM), Yale School of Medicine—Yale Child Study Center (Yale). For detailed acquisition parameters for each site in the ABIDE I cohort, see Table S2.

Data from thirteen different sites were used in the analysis from the ABIDE II cohort: Erasmus University Medical Center (EMC), Rotterdam, ETH Zürich (ETH), Georgetown University (GU), Indiana University [IU], Kennedy Krieger Institute (KKI), Katholieke Universiteit Leuven (KUL), NYU Langone Medical Center Sample 1 (NYU1), NYU Langone Medical Center Sample 2 (NYU2), San Diego State University (SDSU), Stanford University (Stanford), Trinity Centre for Health Sciences (TCD), University of California David (UCD), University of Utah School of Medicine (USM), University of Miami (Miami). For detailed acquisition parameters for each site in the ABIDE II cohort, see Table S3.

All rs-fMRI images were preprocessed using FMRIB Software Library (FSL; www.fmrib.ox.ac.uk/fsl) (10). We applied the following pipeline: removal of the first five volumes to allow for signal equilibration, volume realignment to the middle volume to correct for primary head motion using MCFLIRT, grand mean scaling, and spatial smoothing with a 6mm FWHM Gaussian kernel. We subsequently used ICA-AROMA to correct for secondary head motion-related artifacts (11). ICA-AROMA can remove motion-related artifacts with high accuracy while preserving the neurobiological signal of interest (12) and compares favorably to other methods for removing motion-related confounds (5). Next, we regressed out the mean signals from the CSF and white matter as nuisance covariates and applied a 0.01Hz temporal high-pass filter. The functional images of each participant were co-registered to the participants' anatomical images via boundary-based registration implemented in FSL FLIRT (4, 13). The high-resolution structural images were registered to MNI152 standard space using 12-parameter affine transformation and further refined using a non-linear registration with FSL FNIRT with a 10mm warp and 2mm resampling resolution (10) We finally normalized all the functional images to 2mm MNI152 standard space by applying the transformation of the functional image to T1 and T1 to MNI152. All subsequent analyses were conducted in MNI152 standard space.

There is no current consensus of a preferred pipeline for the analysis of FC case-control differences in autism; therefore, we additionally present results, including global signal regression (GSR), see Figure S5. GSR is a procedure in which the mean signal of all voxels is regressed from each functional image, and it can assist with signal denoising in some circumstances(8) but may affect group differences (14, 15).

**Figure S1. ROIs excluded due to low coverage**

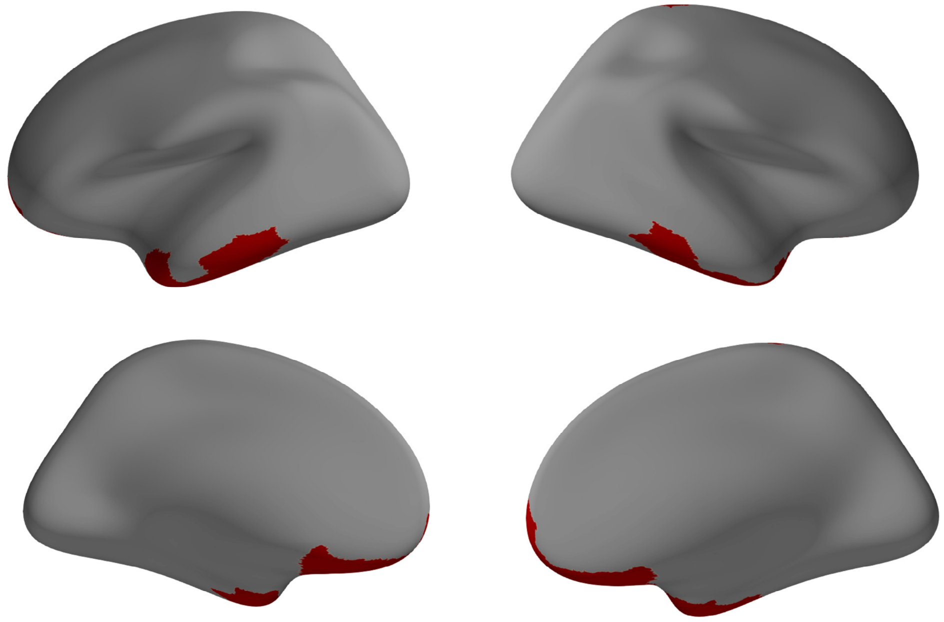

ROIs were excluded (red) for having low coverage (<70% of the voxels in the ROI) in more than 5% of the participants.

### Section 4. Removal of site-specific artifacts

To remove the effects of scanning site, we used ComBat (16), a harmonization technique based on multivariate linear mixed-effects regression and empirical Bayes. This method has previously been shown to eliminate site effects in multi-site resting-state fMRI (17). For the functional connectivity analysis, we applied ComBat while retaining variance specific to the diagnostic group, age, and sex.

For the canonical correlation analysis (CCA) relating functional connectivity disturbances to symptoms, we applied ComBat to the subset of participants with both imaging and symptom data, retaining variance for all of the behavioral variables included in this analysis in addition to age and sex. Figure S3 shows the edges that show significant effects of scanning site before and after ComBat harmonization, indicating that ComBat successfully removed effects of scanning site.

To further test the effects of our harmonization procedure, we trained a one vs. one, multilabel, support vector machine (SVM) linear classifier to classify data by scanning site. We trained the classifier on 70% of the functional connectivity data and tested it on the remaining 30% of the data. Figure S4 shows the ROC AUC across sites before and after ComBat, again demonstrating that ComBat successfully removed the effects of scanning site.

**Figure S2. Functional connectivity edges showing significant effects of scanning site before and absence of significant effects of scanning site after ComBat harmonization**

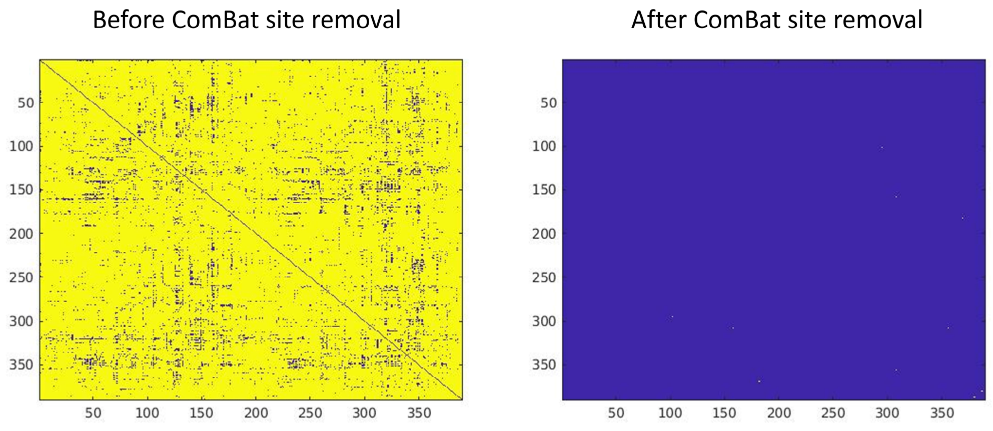

$p<0.05$ in yellow $p>0.05$ in blue

**Figure S3. ROC AUC scores for one vs. one SVM linear classification of scanning sites before and after ComBat**

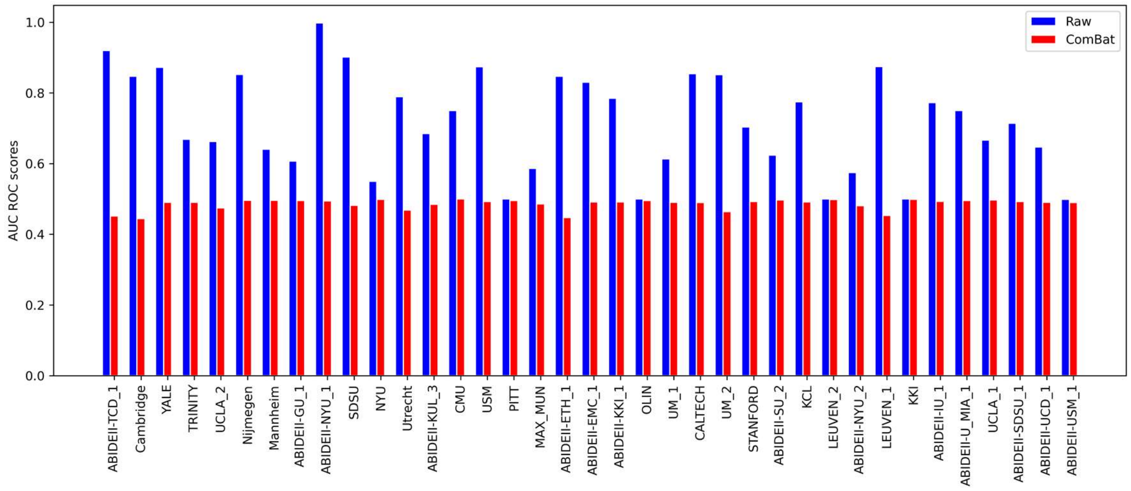

The figure shows the Area Under the Receiver Operating Characteristic Curve (AUC ROC) scores across scanning sites, before and after ComBat.

**Section 5. Mixture modeling normalization**

In this procedure, each correlation value was first normalized using a Fisher-z transformation. Subsequently, three parametrized distributions were fitted, one central Gaussian to model the noise along with two Gamma distributions fitted to the positive and negative tails to model the signal. Next, the connectivity values were normalized by subtracting the mean and dividing by the standard deviation of the obtained Gaussian distribution fitted to the noise (18). This procedure acts like a soft threshold on the data to better separate true from spurious correlations.

**Section 6. Assessing motion-artifact removal**

A commonly used approach to assess whether motion artifacts were successfully removed from the data is to estimate the relationship between the relative RMS displacement (mFD) and the outcome of interest, which in our case is functional connectivity. In order to estimate this relationship after denoising, we computed quality control-functional connectivity (QC-FC) correlations (5, 19).

In scanner head motion is hypothesized to bias connectivity estimates between two nodes in a manner that is related to the distance between those nodes (19-21). This finding is of special importance for neurodevelopmental cohorts, where increased in-scanner motion is a feature distinguishing patients from controls (21). Subject movement may enhance short-distance connections while reducing long-distance connections (19-21). To determine the residual distance dependence of subject movement after denoising for two pipelines, we calculated QC-FC distance dependence (5). We first used the center of mass of each node to obtain the Euclidean distance between the centers of mass of nodes *i* and *j*. We subsequently calculated the correlation between the distance separating each pair of nodes and the *QC-FC* correlation of the edge connecting those nodes; this correlation served as an estimate of the distance-dependence of motion artifact.

Figure S2 shows the QC-FC distributions for our main and alternative pipelines and distance dependence for assessing residual motion contamination in our sample. All QC-FC values were in the range $-0.3 < r < 0.3$, with 95% of the values falling within $-0.15 < r < 0.16$, indicating that residual correlations between FC and head motion were small. These values compare favorably to past benchmarking studies of pipeline denoising efficacy, where some of the best denoising pipelines have QC-FC values varying between $-0.45 < r < 0.45$ (5). Similarly, while some QC-FC distance dependence is still observed, the relationship is consistent with prior reports (5), highlighting the relative success of our denoising approach.

**Figure S4. QC-FC distributions and distance dependence**

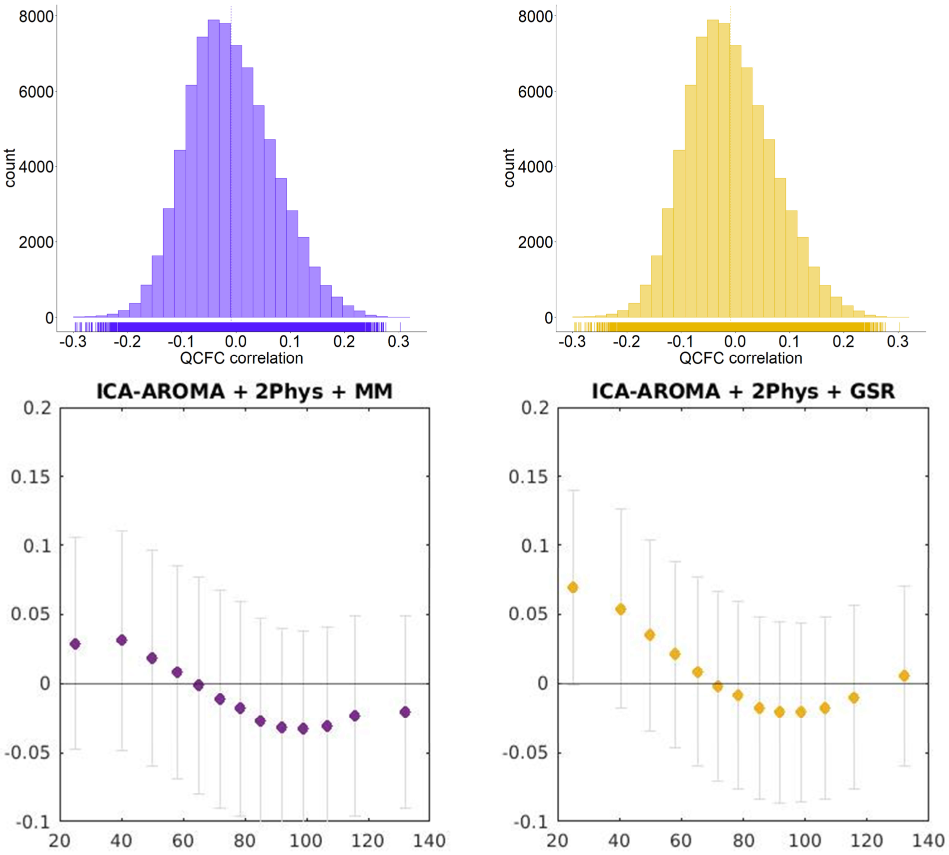

Dependence of the correlation between the framewise displacement and functional connectivity (QC-FC) on the distance between the ROIs. QC-FC correlations were split into 13 equiprobable bins based on the distance between nodes. For each bin, the mean QC-FC for ROI distance bin (circle) and standard deviation (error bar) of QC-FC correlation are shown.

`

### Table S1. Scanning parameters for each scanning site in the EU-AIMS LEAP consortium

| | | | | EPI BOLD multi-echo imaging sequence | | | |
|---|---|---|---|---|---|---|---|
| **Site** | Scanner | Field str. | Instruct. | TR /TE1/TE2 / TE3 (ms)/FA (°) | No. of vol. | No. of slices | Voxel size |
| **Cambridge** | Siemens Magnetom Verio | 3 T | Fixation | 2300/ 12/29/46/80 | 266 | 33 | 3.8 x 3.8 x 3.8 |
| **KCL** | GE NA | 3 T | Fixation | 2300/ NA/31/48/90 | 215 | 33 | 3.8 x 3.8 x 3.8 |
| **Mannheim** | Siemens Magnetom TIM Trio | 3 T | Fixation | 2300/ 12/29/46/80 | 215 | 33 | 3.8 x 3.8 x 3.8 |
| **Nijmegen** | Siemens Magnetom Skyra | 3 T | Fixation | 2300/ 12/NA/NA/80 | 266 | 33 | 3.8 x 3.8 x 3.8 |
| **Utrecht** | Philips | 3 T | Fixation | 2300/ 13/31/49/80 | 200 | 33 | 3.75 x 3.75 x 3.75 |

### Table S2. Scanning parameters for each scanning site in the ABIDE 1 cohort

| | | | | EPI BOLD imaging sequence | | | |
|---|---|---|---|---|---|---|---|
| **Site** | Scanner | Field str. | Instruct. | TR /TE (ms)/FA (°) | No. of vol. | No. of slices | Voxel size |
| **Caltech** | Siemens Magnetom Verio | 3 T | Eyes closed | 2000/30/75 | 150 | 34 | 3.5 x 3.5 x 3.5 |
| **CMU** | Siemens Magnetom Verio | 3 T | Eyes closed | 2000/30/73 | 240 | 28 | 3.0 x 3.0 x 3.0 |
| **KKI** | Philips Achieva | 3 T | Fixation | 2500/30/75 | 156 | 47 | 3.0 x 3.0 x 3.0 |
| **Leuven** | Philips | 3 T | Fixation | 1667/33/90 | 250 | 32 | 3.6 x 3.6 x 4.0 |
| **MaxMun** | Siemens Magnetom Verio | 3 T | Fixation | 3000/30/80 | 120/200 | 40 | 3.0 x 3.0 x 3.0 |
| **NYU** | Siemens Magnetom Allegra | 3 T | Fixation | 2000/15/90 | 180 | 33 | 3.0 x 3.0 x 4.0 |
| **Olin** | Siemens Magnetom Allegra | 3 T | Fixation | 1500/27/60 | 210 | 29 | 3.4 x 3.4 x 4.0 |
| **Pitt** | Siemens Magnetom Allegra | 3 T | Eyes closed | 1500/25/70 | 200 | 29 | 3.1 x 3.1 x 4.0 |
| **SBL** | Philips Intera | 3 T | Eyes closed | 2200/30/80 | 200 | 38 | 2.75 x 2.75 x 2.72 |
| **SDSU** | GE MR750 | 3 T | Fixation | 2000/30/90 | 180 | 41 | 3.4 x 3.4 x 3.4 |
| **Stanford** | GE SIGNA | 3 T | Eyes closed | 2000/30/80 | 180 | 29 | 3.1 x 3.1 x 4.5 |
| **Trinity** | Philips Achieva | 3 T | Eyes closed | 2000/28/90 | 150 | 38 | 3.0 x 3.0 x 3.5 |
| **UCLA** | Siemens Magnetom TIM Trio | 3 T | Fixation | 3000/28/90 | 120 | 34 | 3.0 x 3.0 x 4.0 |
| **UM** | GE SIGNA | 3 T | Fixation | 2000/30/90 | 300 | 40 | 3.44 x 3.44 x 3.0 |
| **USM** | Siemens Magnetom TIM Trio | 3 T | Eyes open | 2000/28/90 | 240 | 40 | 3.4 x 3.4 x 3.0 |
| **Yale** | Siemens Magnetom TIM Trio | 3 T | Eyes open | 2000/25/60 | 200 | 34 | 3.4 x 3.4 x 4.0 |

**Table S3. Scanning parameters for each scanning site in the ABIDE 2 cohort**

| | | | | EPI BOLD imaging sequence | | | |
|---|---|---|---|---|---|---|---|
| **Site** | Scanner | Field str. | Instruct. | TR /TE (ms)/FA (°) | No. of vol. | No. of slices | Voxel size |
| **EMC** | GE MR750 | 3 T | Eyes closed | 2000/30/85 | 160 | 37 | 3.6 x 3.6 x 4.0 |
| **ETH** | Philips Achieva | 3 T | Fixation | 2000/25/90 | 210 | 40 | 3.0 x 3.1 x 3.0 |
| **GU** | Siemens Magnetom TIM Trio | 3 T | Eyes open | 2000/30/90 | 154 | 43 | 3.0 x 3.0 x 2.5 |
| **IU** | Siemens Magnetom TIM Trio | 3 T | Eyes open | 813/28/60 | 433 | 42 | 3.4 x 3.4 x 3.4 |
| **KKI** | Philips Achieva | 3 T | Fixation | 2500/30/75 | 156 | 47 | 3.0 x 3.0 x 3.0 |
| **KUL** | Philips | 3 T | Fixation | 2500/30/90 | 162 | 45 | 2.5 x 2.5 x 2.7 |
| **NYU1** | Siemens Magnetom Allegra | 3 T | Fixation | 2000/15/90 | 180 | 33 | 3.0 x 3.0 x 4.0 |
| **NYU2** | Siemens Magnetom Allegra | 3 T | Fixation | 2000/30/82 | 180 | 34 | 3.0 x 3.0 x 3.0 |
| **SDSU** | GE MR750 | 3 T | Fixation | 2000/30/90 | 180 | 41 | 3.4 x 3.4 x 3.4 |
| **Stanford** | GE SIGNA | 3 T | Eyes closed | 2000/30/80 | 180 | 31 | 3.4 x 3.4 x 3.5 |
| **TCD** | Philips Achieva | 3 T | Fixation | 2000/27/90 | 210 | 37 | 3.0 x 3.0 x 3.2 |
| **UCD** | Siemens Magnetom TIM Trio | 3 T | Eyes open | 2000/24/90 | 460 | 36 | 3.5 x 3.5 x 4.0 |
| **USM** | Siemens Magnetom TIM Trio | 3 T | Eyes open | 2000/28/90 | 240 | 40 | 3.4 x 3.4 x 3.0 |
| **Miami** | GE Healthcare | 3 T | Eyes closed | 2000/30/75 | 290 | NA | 3.4 x 3.4 x 3.0 |

**Section 7.** The Schaefer parcellation and the Network based statistic (NBS)

We used the Schaefer parcellation to divide the cortex of each participant into 400 functional regions of interest (ROIs). The Schaefer parcellation has been demonstrated to generalize well in participants ranging from 6 to 85, which makes it well suited for studies across the human lifespan (22). Furthermore, this parcellation showed higher parcel homogeneity compared to four other well-known parcellations and has a relatively small parcel size that limits problems caused by variations in ROI size (22).We mapped connectome-wide differences between autism and healthy individuals using the Network Based Statistic (NBS)(23), a commonly-used, permutation-based network analog to the cluster-based inference used in typical voxel-wise mapping studies (software available at www.nitrc.org/projects/nbs/). The network based statistic (NBS) addresses the multiple comparison problem inherent in connectome-wide analyses by performing statistical inference at the level of connected components of edges (23). A connected component is a set of nodes that can be linked by a path of edges. Thus, for each contrast, we estimated a test statistic at each edge and applied a primary component-forming threshold, $\tau$, to the data. The size of each connected component in the thresholded matrix was then estimated, and the process repeated 5000 times after shuffling patient and control labels. Following each permutation, the maximal component size was stored to generate an empirical null distribution of maximum component sizes obtained under random group assignment. This null distribution was then used to estimate the statistical significance of the observed component size. Since the null distribution is generated using the maximal statistic, the resulting p-value controls the family-wise error rate at the nominal level (alpha=0.05) and reflects inference performed at the level of connected subnetworks of edges rather than individual connections.

The choice of a primary component forming threshold, $\tau$, is arbitrary. Liberal thresholds are sensitive to weaker yet distributed effects; more stringent thresholds are sensitive to stronger effects with a potentially more focal distribution across the connectome. Here, we follow prior work (24) and report results using *t*-statistic thresholds of $\tau$ = {1.7, 2.3, and 3.1}, corresponding to p = {.05, .01 and .001} uncorrected. Note that the $\tau$ threshold is distinct from the component-wide threshold used to identify significant NBS components, which does control the familywise error.

**Section 8. Behavioral measures**

The behavioral measures that we used in our analysis comprised the social affect, communication, and restrictive and repetitive behaviors subscales of the Autism Diagnostic Observation Schedule calibrated severity score (ADOS); the social domain, communication domain, and restricted and repetitive behaviors domain scores of the Autism Diagnostic Interview-Revised (ADI-R); the Social Responsiveness Scale second edition (SRS-2) raw scores(25); the daily living skills, socialization, and communication subscales of the Vineland Adaptive Behavior Scale (VABS)(26) and the Short Sensory Profile subscales (27).

While the ADI-R investigates the developmental history of an individual through an interview, conducted by a professional, with the parents or caregivers (28), the ADOS provides information with respect to the individuals' current symptoms during a semi-structured interaction with an examiner (29, 30) ADOS 2 scores from the ABIDE datasets were converted to ADOS calibrated severity scores (31, 32) to match the scores from EU-AIMS LEAP.

The SRS-2 assesses the presence and severity of social impairment in autism. The SRS-2 assessment is in the form of parent and/or teacher ratings for individuals younger than 18 and self-report for individuals aged 18 and higher (25). The VABS is a standardized assessment tool that supports the diagnosis of intellectual and developmental disabilities by the use of a semi-structured interview measuring adaptive behavior in daily living activities, communication, and social relationships (26).

The Short Sensory Profile (SSP) is a shortened form of Dunn's Sensory Profile caregiver questionnaire (33) originally developed for screening and identification of children with sensory processing difficulties (27). The SSP includes 38 items organized into seven subscales: Movement Sensitivity (3 items), Tactile Sensitivity (7 items), Sensation Seeking/ Under responsiveness (7 items), Visual/Auditory Sensitivity (5 items), Low Energy/Weak (6 items), Taste/Smell Sensitivity (4 items), and Auditory Filtering (6 items). Since lower scores of the SSP subscales indicate higher symptom severity, we inverted the scores to correspond to lower scores indicating lower symptom severity. We performed this procedure in order to ensure better clarity and readability of the results. We excluded the auditory filtering subscale of the SSP as it was strongly correlated ($r>0.7$) with two of the other subscales.

**Section 9. Results from global signal regression (GSR) pipeline**

The general pattern of findings was consistent across pipelines (Figure S5). The main difference in the two pipelines' findings was the connectivity between the visual and somatomotor network, which in the ICA-AROMA + GSR pipeline was found in the hyperconnectivity network. In contrast, in the ICA-AROMA + MM pipeline, it was included in the hypoconnectivity network.

**Figure S5. Case-control differences in functional connectivity for the main preprocessing pipeline (ICA-AROMA + MM) and the alternative pipeline (ICA-AROMA + GSR).**

ICA-AROMA + MM

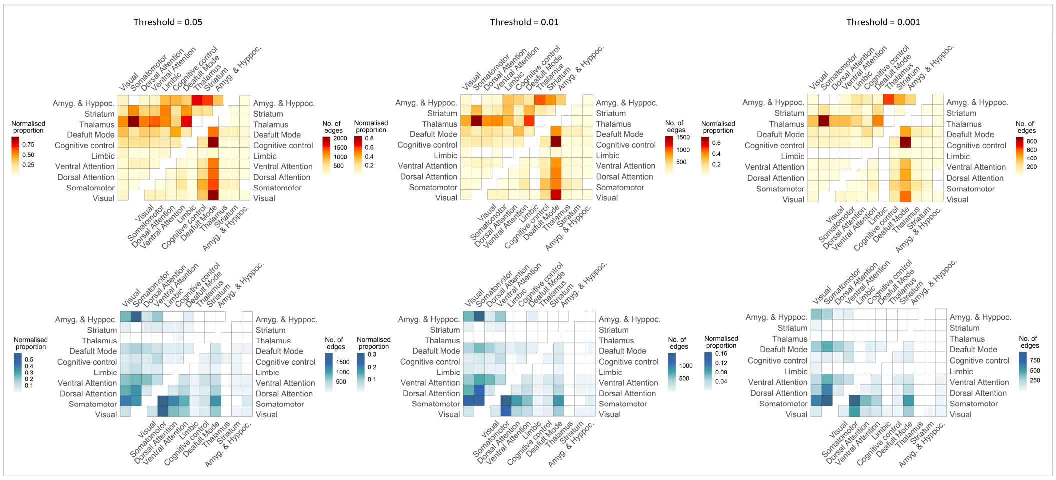

ICA-AROMA + GSR

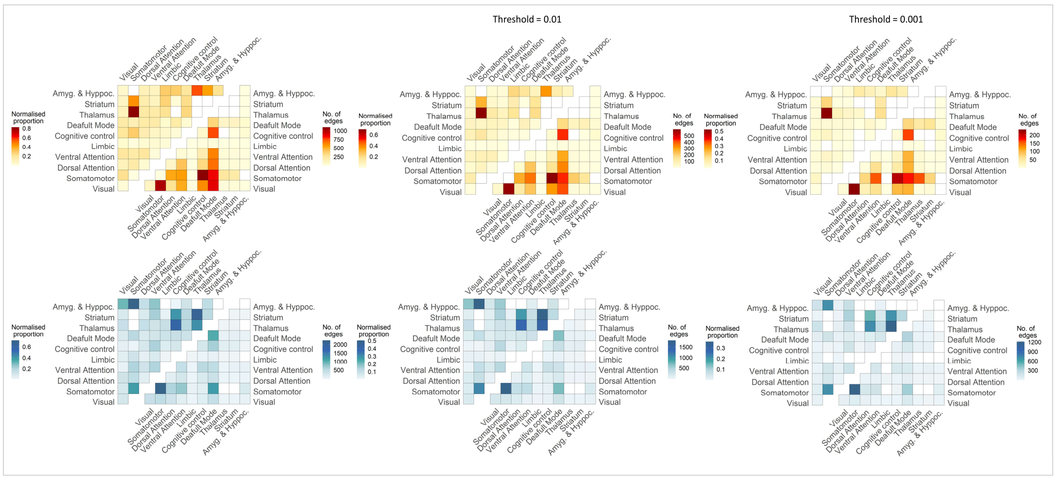

### Section 10. Sensitivity analyses

*IQ*

To check the consistency of our findings we performed an additional analysis in which we included full-scale Intelligence quotient (IQ) as a covariate in the model. Full-scale IQ was available for 1769 subjects, with an age range of 5-58. See Table 6 for a detailed characterization of this subsample. Figure S6 shows that the case-control differences in functional connectivity were almost identical to our primary findings. Interactions between diagnosis and age or sex were not significant.

#### Figure S6. Case-control differences in functional connectivity when covaried for full-scale IQ

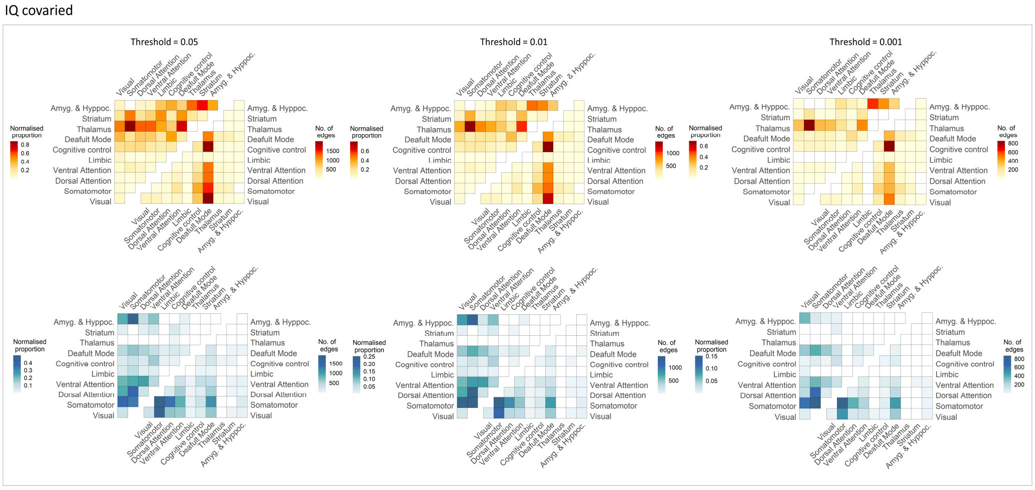

**Table S4. Demographic information of the subsample used for the IQ covaried analysis**

| | Autism | NT | test value, p-value |
|---|---|---|---|
| **n** | 775 | 994 | - |
| **Male/female [a]** | 637/138 | 744/250 | **13.3, 0.0003** |
| **Age (mean ± SD)** | 16.7 ±7.2 | 16.8 ±7.0 | 1.7, 0.09 |
| **Full-scale IQ (mean ± SD, n)** | 106.1 ±15.8, 775 | 112.2 ±12.8, 994 | **-8,913, p<0.0001** |
| **Head motion meanFD (mean ± SD)** | 0.079 ±0.037 | 0.072 ±0.033 | **4.3, p<0.0001** |
| **Handedness (right/left/ambidextrous, n)** | 505/70/31, 606 | 727/50/24, 801 | - |
| **Current medication use** | 198 | 21 | - |

*Analysis including subjects with IQ<70*

In our main analysis, we excluded subjects with IQ<70, because of concerns of the multiple comorbidities causing intellectual disability in the control group subjects with IQ<70. Here we repeat our main analysis to include also the subjects with IQ<70. Figure S10 shows that the case-control differences in functional connectivity were consistent with our primary findings; thus, our conclusions remain unchanged.

**Figure S7. Case-control differences in functional connectivity of a sample additionally including subjects with IQ<70**

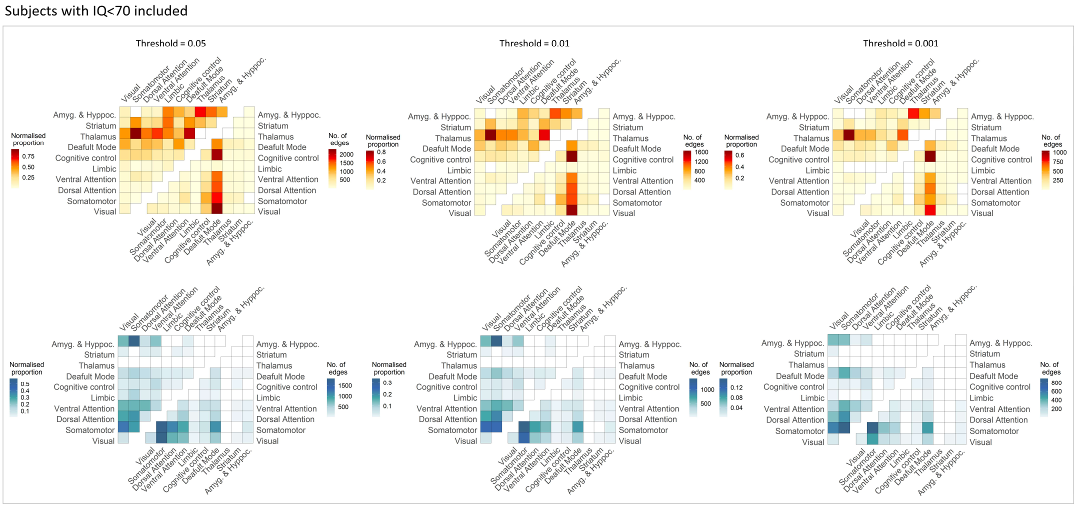

**Table S5. Demographic information of the sample additionally including subjects with IQ<70**

| | Autism | NT | test value, p-value |
|---|---|---|---|
| **n** | 829 | 1046 | - |
| **Male/female [a]** | 678/151 | 780/266 | **13.5, p=0.0002** |
| **Age (mean ± SD)** | 16.6 ±7.1 | 16.0 ±6.9 | 1.95, p=0.06 |
| **Full-scale IQ (mean ± SD, n)** | 104.5 ±17.3, 808 | 111.3 ±14.3, 1012 | **-9.1, p<0.0001** |
| **Head motion meanFD (mean ± SD)** | 0.12 ±0.28 | 0.08 ±0.14 | **3.3, p=0.0008** |
| **Handedness (right/left/ambidextrous, n)** | 542/81/31, 654 | 763/57/24, 844 | - |
| **Current medication use** | 205 | 21 | - |

*ADHD comorbidity*

Attention deficit hyperactivity disorder (ADHD) is the most common comorbidity of ASD (34, 35). In addition to the behavioral, biological, and neuropsychological overlap between the two disorders, there are also important differences (35) that could potentially influence our results.

To ensure that our findings are not driven by ADHD-specific functional connectivity, we performed an additional analysis, in which we excluded subjects with ADHD comorbidity (N=155). See Table 7 for a detailed characterization of this subsample. Figure S7 shows that the case-control differences in functional connectivity were consistent with our primary findings; thus, our conclusions remain unchanged. Interactions between diagnosis and age or sex were not significant.

### Figure S8. Case-control differences in functional connectivity of the subsample excluding subjects with ADHD comorbidity

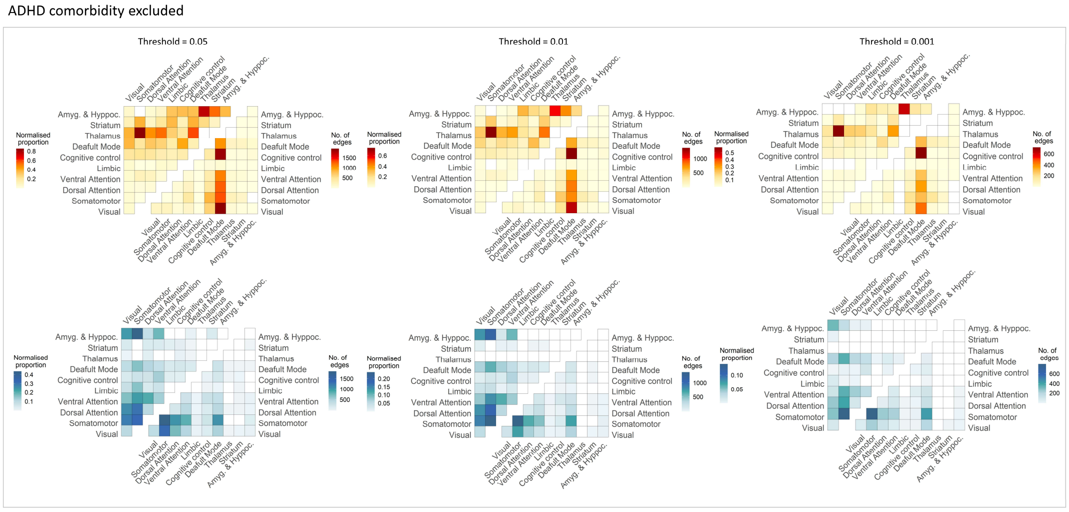

### Table S6. Demographic information of the subsample excluding subjects with ADHD comorbidity

| | Autism | NT | test value, p-value |
|---|---|---|---|
| **n** | 651 | 1018 | - |
| **Male/female** [a] | 546/105 | 768/250 | **16.4, p<0.0001** |
| **Age (mean ± SD)** | 16.9 ±7.4 | 15.9 ±6.9 | **2.8, p=0.006** |
| **Full-scale IQ (mean ± SD, n)** | 106.6 ±15.7, 633 | 112.3 ±12.7, 984 | **-8.0, p<0.0001** |
| **Head motion meanFD (mean ± SD)** | 0.077 ±0.035 | 0.072 ±0.033 | **3.0, p<0.0001** |
| **Handedness (right/left/ambidextrous, n)** | 424/56/22, 502 | 744/56/24, 824 | - |
| **Current medication use** | 149 | 19 | - |

Sex

To ensure that our findings are not sensitive to the imbalanced males to female ratio in our sample, we performed an additional analysis, in which we excluded all 397 female subjects. See Table 8 for a detailed characterization of this subsample. Figure S8 shows that the case-control differences in functional connectivity were consistent with our primary findings; thus, our conclusions remain unchanged.

### Figure S9. Case-control differences in functional connectivity of the subsample including only male subjects

Only male subjects

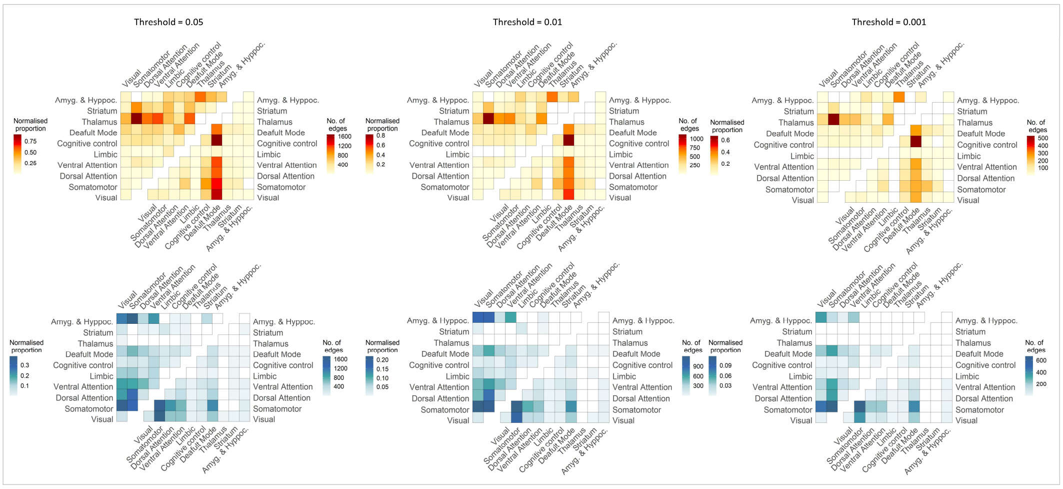

### Table S7. Demographic information of the subsample including only male subjects

| | Autism | NT | test value, p-value |
|---|---|---|---|
| **n** | 655 | 772 | - |
| **Male/female** [a] | 655/0 | 772/0 | - |
| **Age (mean ± SD)** | 16.4 ±.7.0 | 16.3 ±7.1 | **0.1, p=0.9** |
| **Full-scale IQ (mean ± SD, n)** | 106.4 ±16.1, 637 | 112.2 ±12.6, 744 | **-7.4, p<0.0001** |
| **Head motion meanFD (mean ± SD)** | 0.08 ±0.036 | 0.073 ±0.033 | **3.6, p=0.0003** |
| **Handedness (right/left/ambidextrous, n)** | 418/61/26, 505 | 554/43/18, 615 | - |
| **Current medication use** | 163 | 15 | - |

*Medication*

To ensure that the use of psychoactive medication, which is relatively common in individuals with autism, did not affect our results, we performed an additional analysis, in which we excluded 226 participants that reported psychoactive medication use. See Table 9 for a detailed characterization of this subsample. Figure S8 shows that the case-control differences in functional connectivity were consistent with our primary findings; thus, our conclusions remain unchanged.

### Figure S10. Case-control differences in functional connectivity of the subsample including only unmedicated subjects

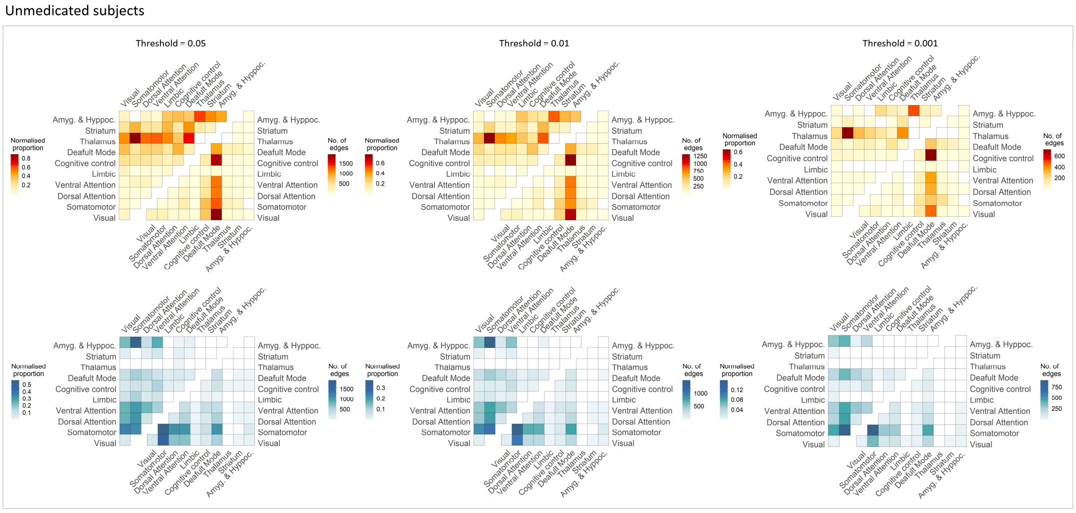

### Table S8. Demographic information of the subsample including only unmedicated subjects

| | Autism | NT | test value, p-value |
|---|---|---|---|
| **n** | 591 | 1007 | - |
| **Male/female** [a] | 492/99 | 757/250 | **13.7, p=0.0002** |
| **Age (mean ± SD)** | 16.7 ±7.5 | 15.9 ±6.9 | **2.1, p=0.04** |
| **Full-scale IQ (mean ± SD, n)** | 106.6 ±15.9, 577 | 112.2 ±12.7, 973 | **-7.0, p<0.0001** |
| **Head motion meanFD (mean ± SD)** | 0.079 ±0.036 | 0.072 ±0.033 | **4.2, p<0.0001** |
| **Handedness (right/left/ambidextrous, n)** | 382/48/27, 457 | 740/54/24, 818 | - |
| **Current medication use** | 0 | 0 | - |

### Section 11. Replication

To further test the robustness of our findings we conducted the connectivity mapping analysis in the LEAP and ABIDE1&2 samples separately. The results we obtained followed a very similar pattern of functional connectivity differences observed in our primary analysis. To quantifu the extent of replication we correlated the network counts of the hypo and hyperconnectivity of the two results (hypoconnectivity: r=0.93,p=0; hyperconnectivity: r=0.82,p=0) and the t-statistics on the level of individual edges (r=0.5; p=0). Thus, the atypical functional connectivity initially observed replicates in two independent samples separately (Figure S11).

#### Figure S11 Case-control differences in functional connectivity replicated in the LEAP sample and ABIDE1&2 samples separately

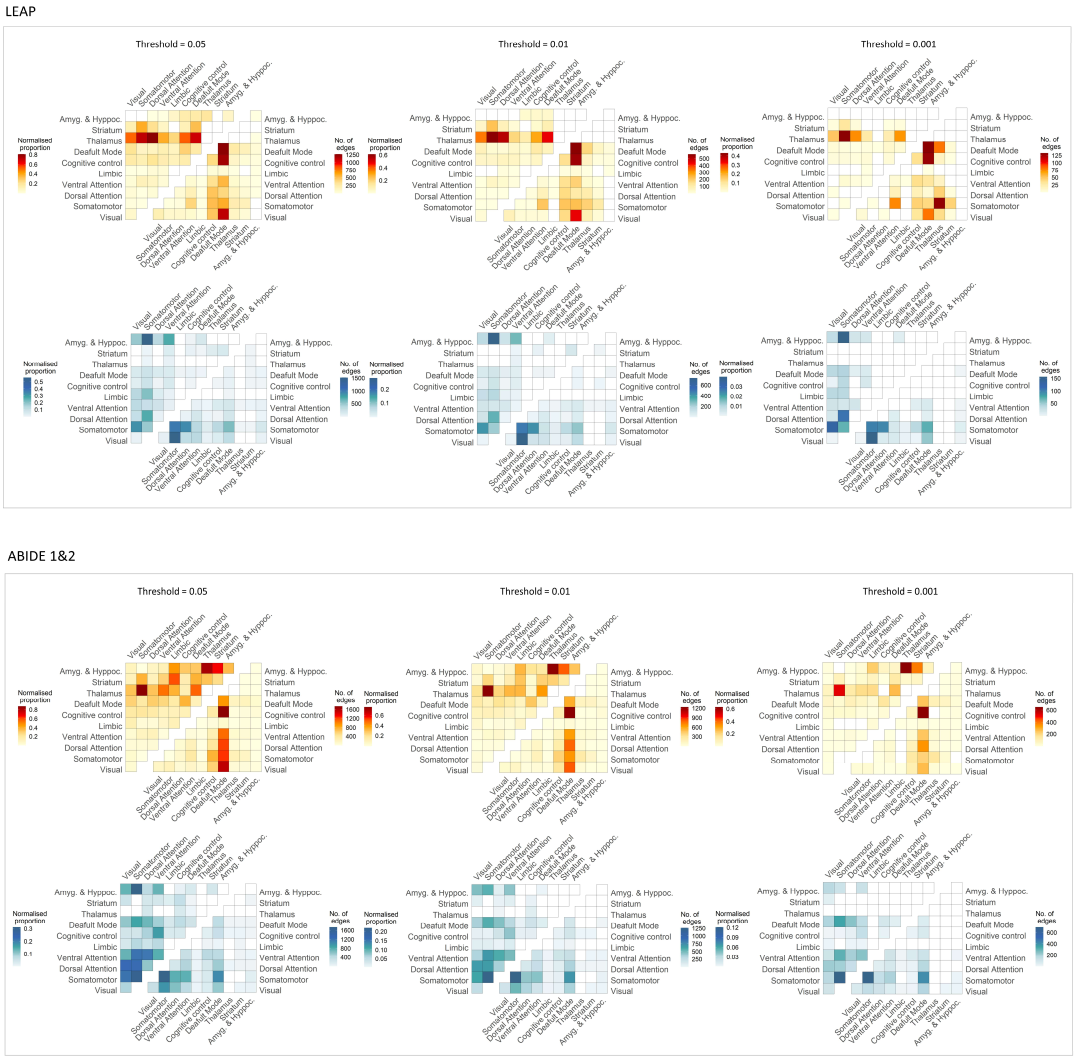

**Section 12. Age by diagnosis interaction**

Our age-by-diagnosis interaction analysis at the ASD>NT contrast revealed a network component ($p_{fwe}$=<0.08) at the primary threshold of $p$=0.05. This component included connections between the visual network and other cortical networks, as well as connections between the cognitive control network and the rest of the cortical networks. The highest proportion of edges implicated in this interaction was observed between the thalamus and the somatomotor, dorsal attention and salience networks as well as between the thalamus and the amygdala & hippocampus. Post-hoc analysis revealed that while the strength of the connections in this network decreased with age in the NT controls, it showed a slight increase with age in the autism sample (Figure S5). It should, however, be noted that this effect failed to reach statistical significance after correction for the two-tailed t-test and should therefore be interpreted with caution.

**Figure S12. Age interaction of the contrast ASD>NT**

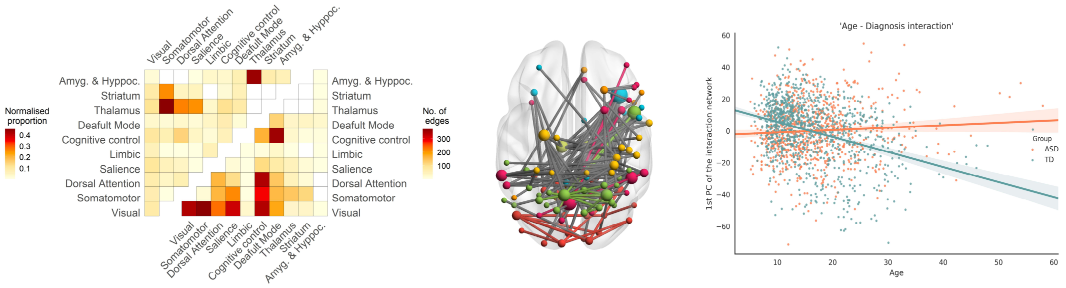

**Section 13. Canonical Correlation Analysis (CCA)**

We used CCA to relate autism-related FC connectivity to symptom severity within the autism group. CCA finds linear mixtures, known as canonical variates (CV), in two sets of variables by maximizing the correlation of the CV of one set with a corresponding CV of the other while being uncorrelated with all other CVs in either set (36). In our analyses, one set of variables included measures of behavior and symptom severity, and the other set of measures included FC estimates for edges identified as showing atypical connectivity in our NBS analysis i.e., the hypo- and hyperconnectivity networks combined.

Since the NBS identified a large number of edges showing FC differences between the autism and control groups, we used Principal Component Analysis (PCA) to reduce the dimensionality of the set of edges showing significant FC differences between the autism and control group, retaining a sufficient number of components to account for

50% of the variance in FC (48 components for CCA1, 44 for CCA2). Age and sex were used as confounds, and non-parametric statistical inference for the CCA was performed using 10 000 permutations(36), with the constraint that permutations were only performed within each study site to account for site effects. To interpret the resulting canonical variates, we evaluated the loadings (also known as structure coefficients), of each individual behavioral variable and FC at each individual edge with the variate scores generated by the CCA.

To further demonstrate the robustness of our findings and show that scanning site does not impact our results in a meaningful way, we applied CCA with leave-one-site-out validation. CCA was applied repeatedly, each time excluding one site. The behavioral score loadings were computed by averaging the loadings across the runs. The FC loadings were obtained by first averaging the U variates across runs and correlating the mean U with the FC at each edge across subjects.We consider as significant only the loadings that were significant across all of the runs for the behavioral variables set. We used 10 000 bootstraps with replacement to estimate the standard error of each loading estimate, allowing us to compute a z-score and associated p-value for each correlation. We identify reliable and statistically significant loadings as those that survive a False Discovery Rate (FDR)-corrected threshold of $p<.05$. See Table S11 and Table S12 for p-values at each fold.

**Figure S13. CCA Leave-one-site analysis results (Social, RRB, adaptive behavior, and IQ)**

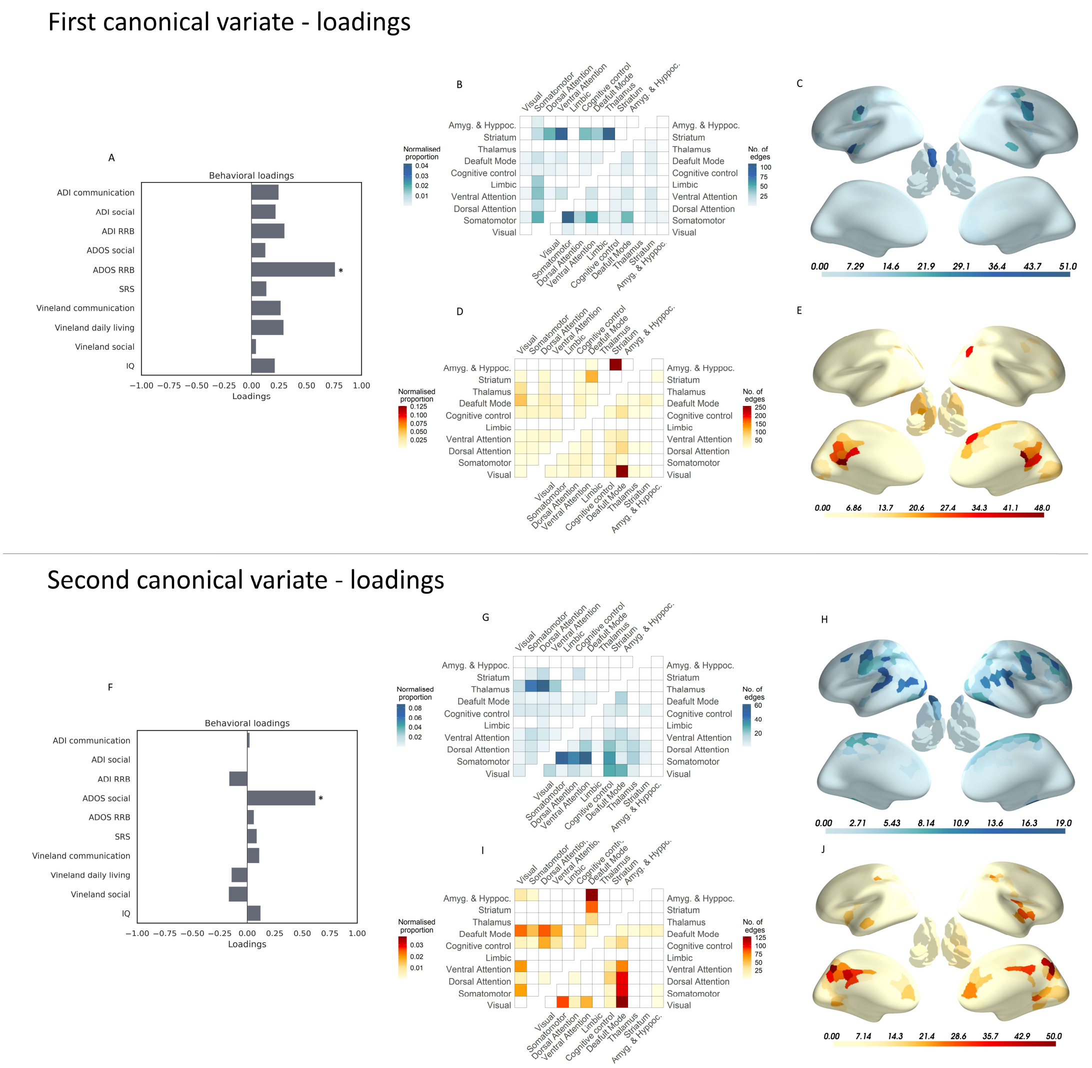

A) Structural coefficients, also known as loadings of the behavioral scores with the first CV; B) Significant negative FC loadings of CV1:p<0.05; C) Degree centrality of the significant negative FC loadings of CV1 D) Significant positive FC loadings of CV1: p<0.05; E) Degree centrality of the significant positive FC loadings of CV1; F) Structural coefficients, also known as loadings of the behavioral scores with the first CV; G) Significant negative FC loadings of CV1:p<0.05; H) Degree centrality of the significant negative FC loadings of CV1; I) Significant positive FC loadings of CV1: p<0.05; J) Degree centrality of the significant positive FC loadings of CV1

**Figure S14. CCA Leave-one-site analysis results (SSP subscales)**

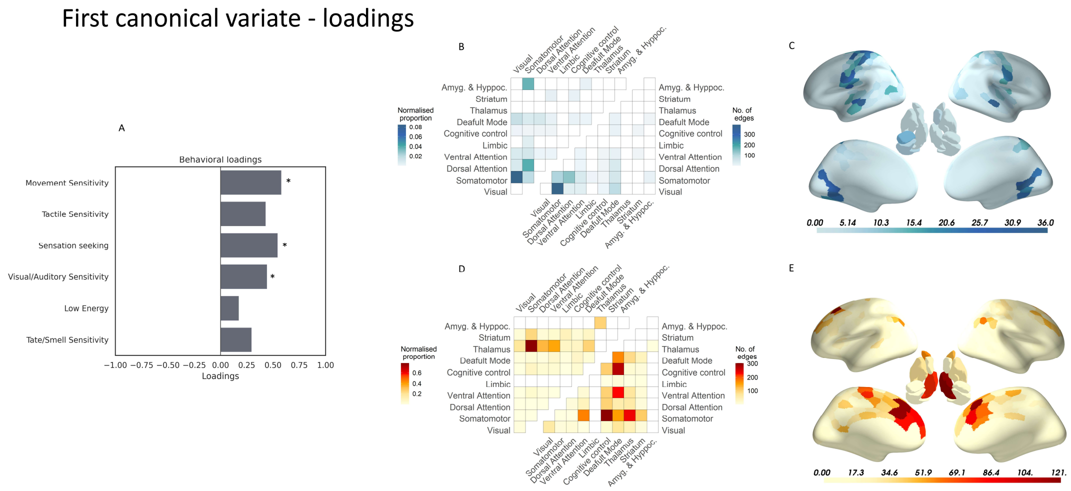

A) Structural coefficients, also known as loadings of the behavioral scores with the first CV; B) Significant negative FC loadings of CV1: $p<0.05$; C) Degree centrality of the significant negative FC loadings of CV1 D) Significant positive FC loadings of CV1: $p<0.05$; E) Degree centrality of the significant positive FC loadings of CV1.

### Table S9. Demographic and clinical information of the subsample used for CCA (Social, RRB, and adaptive behavior)

| | Autism | NT | test value, p-value |
|---|---|---|---|
| **n** | 232 | 0 | - |
| **Male/female [a]** | 171/61 | - | - |
| **Age<br>(mean ± SD)** | 14.8<br>±6.5 | - | -<br>- |
| **Full-scale IQ (mean ± SD, n)** | 104.9 ±16.8, 232 | - | - |
| **Head motion meanFD (mean ± SD)** | 0.081 ±0.04 | - | - |
| **Handedness (right/left/ambidextrous, n)** | 137/24/15, 176 | - | - |
| **Current medication use** | 67 | - | - |
| **ADOS total (mean ± SD, n)** | 5.7 ±2.6, 232 | - | - |
| **ADOS social (mean ± SD, n)** | 6.0 ±2.4, 232 | | |
| **ADOS RRB (mean ± SD, n)** | 5.6 ±2.9, 232 | | |
| **ADI social (mean ± SD, n)** | 16.7 ±6.5, 232 | - | - |
| **ADI communication (mean ± SD, n)** | 14.0 ±5.5, 232 | - | - |
| **ADI RRB (mean ± SD, n)** | 4.5 ±2.6, 232 | - | - |
| **SRS t-score (mean ± SD, n)** | 86.7 ±30.4,232 | - | - |
| **VABS composite (mean ± SD, n)** | 145.6 ± 100.5, 232 | - | - |
| **VABS daily living skills (mean ± SD, n)** | 81.0 ±15.1, 232 | - | - |
| **VABS socialization (mean ± SD, n)** | 76.8 ± 14.3, 232 | - | - |
| **VABS communication (mean ± SD, n)** | 81.4 ±14.3, 232 | - | - |

### Table S10. Demographic and clinical information of the subsample used for CCA (Short Sensory Profile subscales)

| | Autism | NT | test value, p-value |
|---|---|---|---|
| **n** | 125 | 78 | - |
| **Male/female** [a] | 87/38 | 52/26 | 0.07, 0.7 |
| **Age (mean ± SD)** | 16.6 ±5.6 | 14.2 ±3.1 | **3.4, p=0.0007** |
| **Full-scale IQ (mean ± SD, n)** | 105.7 ±15.5, 125 | 109.5 ±11.5, 77 | -1.8, p=0.07 |
| **Head motion meanFD (mean ± SD)** | 0.078 ±0.041 | 0.073 ±0.04 | 0.76, p=0.4 |
| **Handedness (right/left/ambidextrous, n)** | 104/13/2, 119 | 71/5/1, 77 | - |
| **Current medication use** | 45 | 5 | - |
| **SSP movement sensitivity (mean ± SD, n)** | 12.7 ±2.9, 125 | 14.4 ±1.5, 78 | **-4.7, p<0.0001** |
| **SSP tactile sensitivity (mean ± SD, n)** | 27.7 ±5.7,125 | 33.7 ±2.5, 78 | **-8.8, p<0.0001** |
| **SSP sensation seeking (mean ± SD, n)** | 27.3 ±6.4, 125 | 33.3 ±3.0, 78 | **-7.8, p<0.0001** |
| **SSP visual/auditory sensitivity (mean ± SD, n)** | 18.4 ±5.2, 125 | 24.1 ±1.6, 78 | **-9.3, p<0.0001** |
| **SSP low energy (mean ± SD, n)** | 24.4 ±7.0, 125 | 29.3 ±2.2, 78 | **-5.9, p<0.0001** |
| **SSP taste/smell sensitivity (mean ± SD, n)** | 15.5 ±4.7, 125 | 19.1 ±2.1, 78 | **-6.4, p<0.0001** |
| **SSP auditory filtering (mean ± SD, n)** | 17.2 ±5.4, 125 | 26.1 ±3.9, 78 | **-12.4 p<0.0001** |

**Table S11. Canonical correlations and p-values of the significant canonical variates in the leave-one-site-out CCA analyses for the various symptom subscales**

| Site excluded | CV1 | CV2 |
|---|---|---|
| KCL | r = 0.78; p < 0.0001 | r = 0.70; p < 0.0001 |
| Nijmegen | r = 0.74; p < 0.0001 | r = 0.71; p < 0.0001 |
| Utrecht | r = 0.74; p < 0.0001 | r = 0.67; p < 0.0001 |
| Cambridge | r = 0.73; p < 0.0001 | r = 0.66; p < 0.0001 |
| NYU ABIDE 1 | r = 0.75; p < 0.0001 | r = 0.69; p < 0.002 |
| NYU 1 ABIDE 2 | r = 0.73; p < 0.0001 | r = 0.65; p < 0.0001 |
| NYU 2 ABIDE 2 | r = 0.74; p < 0.0001 | r = 0.69; p < 0.0001 |

**Table S12. Canonical correlations and p-values of the significant canonical variates in the leave-one-site-out CCA analyses for the SSP subscales**

| Site excluded | CV1 |
|---|---|
| KCL | r = 0.70; p = 0.008 |
| Nijmegen | r = 0.72; p = 0.03 |
| Utrecht | r = 0.65; p < 0.0001 |
| Mannheim | r = 0.66; p = 0.003 |
| Cambridge | r = 0.67; p = 0.006 |

**Figure S15. Age distribution**

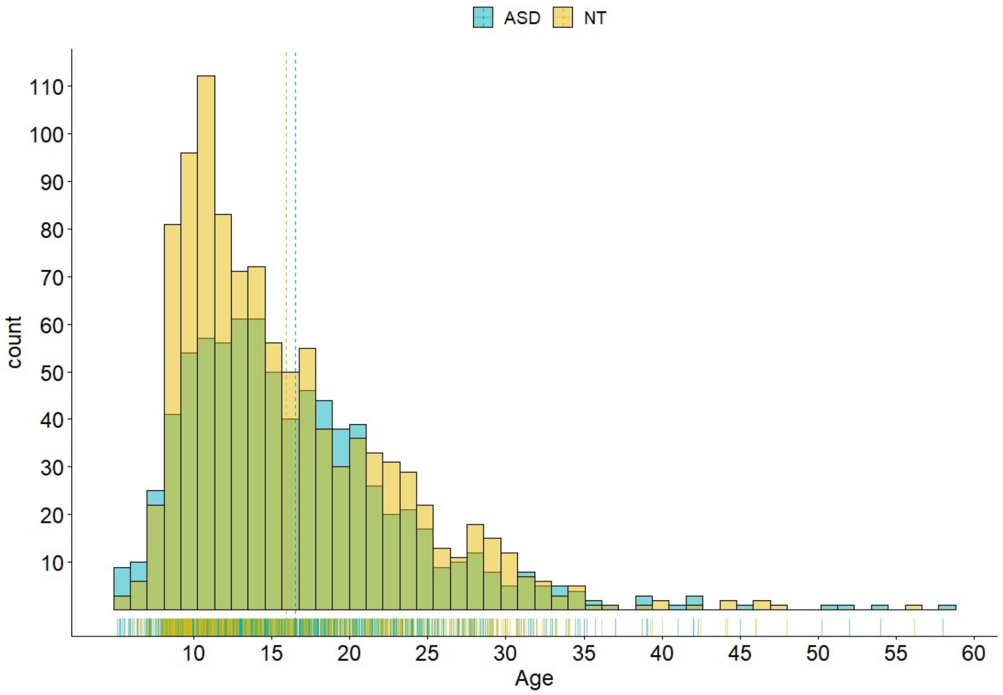

Histograms of the age distributions of ASD and controls
